# A Systematic Exploration of LLM Behavior for EHR Phenotyping

**DOI:** 10.64898/2026.04.16.26350890

**Authors:** Eric Yamga, Shawn Murphy, Philippe Després

**Affiliations:** Department of Medicine, Centre hospitalier de l’Université de Montréal (CHUM), Montréal, Québec, Canada; Centre de recherche du Centre hospitalier de l’Université de Montréal (CRCHUM), Montréal, Québec, Canada; Département de physique, de génie physique et d’optique, Université de Laval, Québec, Québec, Canada; Department of Neurology, Massachusetts General Hospital, Boston, MA; Department of Biomedical Informatics and Medical Education, Seattle, WA

## Abstract

**Background:** Electronic health record (EHR) phenotyping underpins observational research, cohort discovery, and clinical trial screening. Large language models (LLMs) offer new capabilities for extracting phenotypes from unstructured text, but their performance depends on pipeline design choices-including prompting, text segmentation, and aggregation. No systematic framework has previously examined how these parameters shape accuracy and reproducibility.

**Methods:** We evaluated LLM-based phenotyping pipelines using 1,388 discharge summaries across 16 clinical phenotypes.

A full factorial experiment with LLaMA-3B, 8B, and 70B systematically varied three pipeline components: prompting (zero-shot, few-shot, chain-of-thought, extract-then-phenotype), chunking (none, naive, document-based), and aggregation (any-positive, two-vote, majority), yielding 24 configurations per model.

To compare intrinsic model capabilities, biomedical domain-adapted, commercial frontier (LLaMA-405B, GPT-4o, Gemini Flash 2.0), and reasoning-optimized models (DeepSeek-R1) were evaluated under a fixed configuration. Performance was assessed using precision, recall, and macro-F1; secondary analyses examined prediction consistency (Shannon entropy), self-confidence calibration, and the development of a taxonomy of recurrent model errors.

**Results:** Factorial ANOVAs showed that chunking and aggregation were the dominant drivers of performance, whereas the prompting strategy contributed minimally. Configuration effects were stable across model sizes, with no significant Model x Parameter interactions. Phenotype difficulty varied substantially (macro-F1 = 0.40-0.90), yet the highest-performing configuration-whole-document inference without aggregation-was consistent across phenotypes, as confirmed by mixed-effects modeling.

In cross-model comparisons, DeepSeek-R1 achieved the highest macro-F1 (0.89), while LLaMA-70B matched GPT-4o and LLaMA-405B at substantially lower cost. Prediction entropy was low overall and driven primarily by phenotype difficulty rather than prompting or temperature.

Self-confidence calibration was only moderately informative: high-confidence predictions were more accurate, but larger models exhibited systematic overconfidence.

**Conclusions:** LLM performance in EHR phenotyping is governed primarily by input structure and model capacity, not prompt engineering. Simple, document-level inference yields robust performance across diverse phenotypes, providing practical design guidance for LLM-based cohort identification while underscoring the continued need for human oversight for challenging phenotypes.

## 1 Article Manuscript

### 1.1 Introduction

Cohort identification is a fundamental step in any research involving secondary use of electronic health record (EHR) data^1,2^. This rate-limiting step requires integrating heterogeneous —often noisy and incomplete— clinical data to determine whether patients meet the criteria for specific conditions. Accurate phenotyping is essential to ensure the validity of downstream applications, including retrospective observational studies, predictive modeling, and clinical trial enrollment^3–5^. Yet phenotyping at scale remains challenging: existing methods are time-consuming to develop, labor-intensive to maintain, and highly variable in performance across phenotypes and institutions.

#### 1.1.1 Previous Work and Limitations

Large language models (LLMs) have recently emerged as a promising alternative for EHR phenotyping^6^. Their ability to perform complex language-understanding tasks without task-specific training has enabled them to overcome long-standing limitations of traditional biomedical NLP pipelines in a wide range of clinical NLP benchmarks. Early research demonstrated that prompt-engineered LLMs can match or exceed traditional NLP methods for name-entity recognition, information extraction, and document classification^6–8^. Building on this capability, several groups have begun applying LLMs to EHR phenotyping, demonstrating that LLMs can identify clinical conditions directly from unstructured notes^8–11^. Prior work by Wornow et al. established strong zero-shot LLM performance for trial eligibility matching —which is closely related to EHR phenotyping — and highlighted how pipeline structure materially affects outcomes (e.g., note retrieval strategy and chunk selection affected the LLM performance in their study^9^).

Building on this foundation, we go deeper by using a full factorial design to isolate the independent and interactive effects of model architecture and pipeline components (prompting, segmentation, and aggregation) on EHR phenotyping accuracy and robustness. Whereas prior studies have demonstrated feasibility, none have characterized performance change under different structural choices. Our goal is to fill this gap by providing generalizable guidance for constructing reliable LLM-based EHR phenotyping pipelines.

From our review, we classify LLM-based phenotyping approaches into three broad categories: LLM-to-Phenotype Strategy: Direct classification of entire notes or merged documents using an LLM as a binary classifier. This approach is simple and effective, requiring minimal implementation effort ^8^.

Map-Reduce Strategy: Segmentation of clinical notes into chunks, followed by classification of each chunk, and vote-aggregation of the chunk-level predictions^12^. It addresses the challenge of LLMs’ limited context windows in cases where large corpuses are used^13^.

Extract-Then-Phenotype Strategy: This approach involves a two-step process using the LLM for entity extraction, followed by a deterministic rule-based classifier^14,15^.

Across these efforts, a common pattern emerges: LLM performance depends strongly on how the text is segmented, how prompts are structured, and how intermediate predictions are aggregated. Moreover, the stochastic nature of LLM inference introduces nondeterminism, raising concerns for reproducibility^16^.

Prior work has compared several pipeline designs, but these comparisons remain descriptive, evaluating a limited set of predefined configurations without quantifying the independent or interactive effects of each component^8,9^. This limits generalizability and obscures which decisions truly drive performance.

##### Study Rationale and Objectives

To move beyond descriptive evaluations, we conduct a full factorial experimental analysis of LLM behavior in EHR phenotyping. Rather than optimizing a single best-performing pipeline, our objective is to decompose the independent and joint effects of model size, prompting strategy, chunking method, and aggregation rule.

Our primary analyses evaluate classification performance across configurations and use factorial ANOVA to isolate main and interaction effects across phenotypes. Our secondary analyses characterize other aspects of LLM behavior: prediction consistency (Shannon entropy), self-calibration, and qualitative assessment of failure patterns.

Through this systematic exploration, we seek to establish methodological foundations and practical guidance for deploying LLMs robustly and reproducibly in EHR-based clinical research.

### 1.2 Methods

#### 1.2.1 Study Design

In this study, we investigated how design choices in large-language-model (LLM) pipelines affect automated phenotyping from discharge summaries in two parts (see Figure 1).

**Figure 1:**
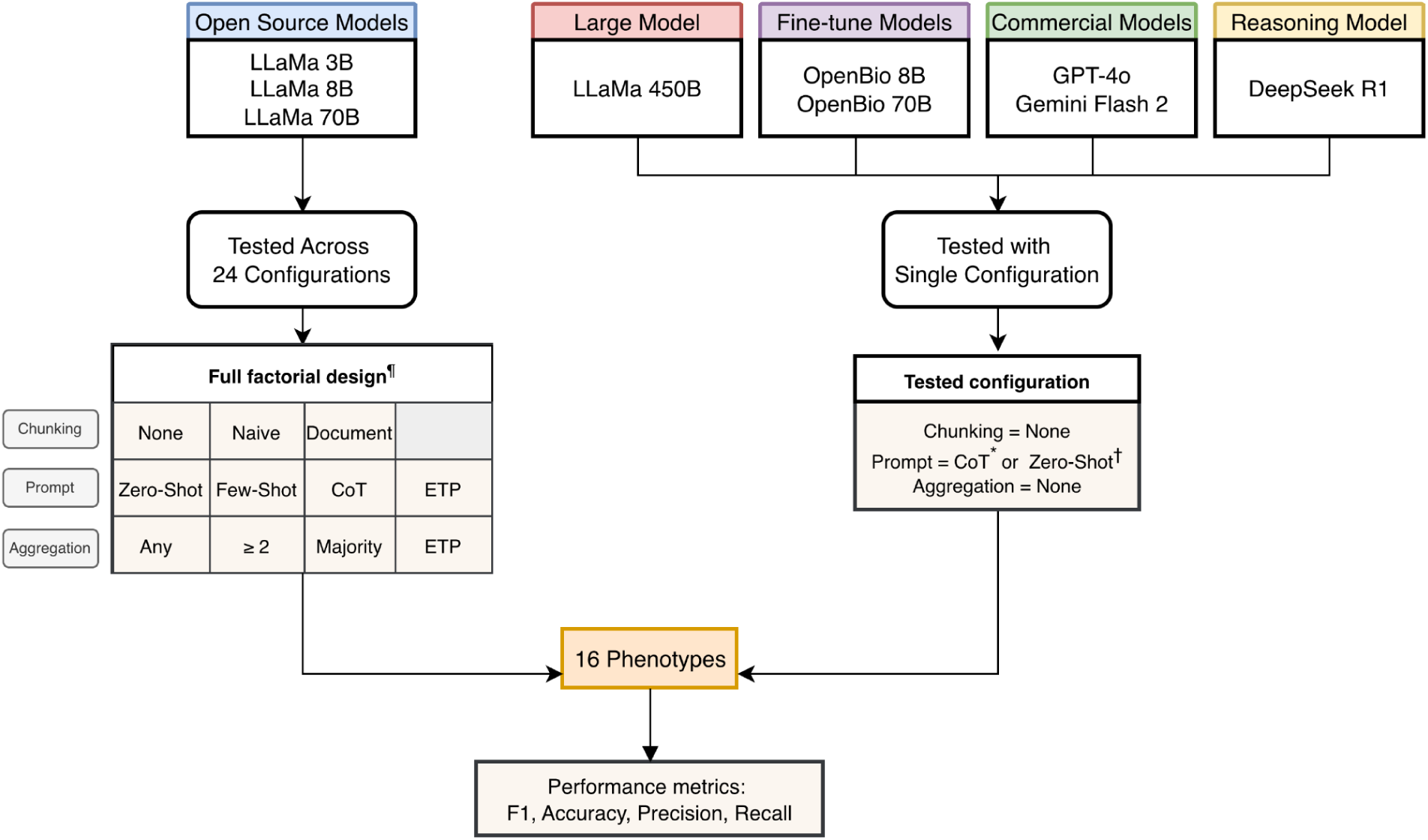
Study flowchart for analyzing large language models behavior in EHR-based phenotyping. Open-source LLaMA models (3B, 8B, 70B) underwent a factorial evaluation across 24 pipeline configurations combining chunking (None, Naïve, Document), prompting (Zero-Shot, Few-Shot, CoT, ETP), and aggregation (Any, > 2, majority voting or the ETP-specific aggregation). In contrast, larger, commercial, and reasoning models were assessed with a fixed configuration (no chunking; †Zero-Shot prompting for DeepSeek, and *CoT prompting for the other models).

First, we conducted a factorial experiment within a single model family across multiple configurations to characterize the contribution of prompt strategy, chunking, and aggregation rules^17^. Second, we performed a cross-model comparison using a fixed pipeline configuration to isolate differences attributable to model class rather than pipeline structure. All experiments were implemented in Python 3.11 using the OpenRouter API for LLM inference (additional technical details are provided in the Supplemental File 1 - Supplementary Methods).

##### Dataset

We used the MIMIC-IV Phenotype Atlas (MIPA), a publicly available benchmark comprising 1,388 de-identified discharge summaries manually reviewed by two clinicians with consensus labels for 16 clinically diverse phenotypes (see Table 1)^18^. Ethical review and informed consent were waived. Labels are openly available; access to raw notes requires Collaborative Institutional Training Initiative (CITI) training and PhysioNet approval.

**Table 1:** Phenotype distribution in the MIPA Dataset (n = 1,388 Discharge Summaries)

| Phenotype | Total cases | Prevalence |
| --- | --- | --- |
| **High Prevalence** |  |  |
| Hypertension (essential) | 940 | 67.70% |
| Depression | 679 | 48.90% |
| Type II Diabetes | 393 | 28.30% |
| **Low Prevalence** |  |  |
| Heart Failure (preserved ejection fraction) | 217 | 15.60% |
| Metastatic Cancer | 168 | 12.10% |
| Rheumatoid Arthritis | 164 | 11.80% |
| Heart Failure (reduced ejection fraction) | 122 | 8.80% |
| Systemic Lupus | 113 | 8.10% |
| Dementia | 102 | 7.30% |
| **Lifestyle-Associated Factors** |  |  |
| Obesity | 286 | 20.60% |
| Alcohol Abuse | 155 | 11.20% |
| **Healthcare-Associated Events** |  |  |
| DVT/PE (past history) | 312 | 22.50% |
| DVT/PE (complication) | 39 | 2.80% |
| C. difficile (past history) | 101 | 7.30% |
| C. difficile (complication) | 42 | 3.00% |
| **Other** |  |  |
| None | 35 | 2.50% |

*Acronyms:* DVT (Deep Vein Thrombosis); PE (Pulmonary Embolism).

##### Models

###### Factorial Experiment

For the factorial experiment we selected the open-source models LLaMa 3 at three scale sizes: 3B, 8B, and 70B. Holding the family constant allows to isolate the effect of model size rather than architecture, improving the internal validity of the experiment.

###### Cross-Model Comparisons

For cross-model comparisons, we evaluated LLaMA-3 405B (in addition to the smaller LLaMA-3 models), two biomedical domain–adapted LLaMA-3 variants (OpenBioLLM - 8B, 70B^19^), two commercial frontier models (GPT-4o^20^ and Gemini Flash 2.0), and one reasoning-optimized model (DeepSeek-R1^21^).

OpenBioLLM models were selected because, at the time of study conception, they represented the best-performing publicly available healthcare-focused fine-tuned models. As direct adaptations of the LLaMA-3 architecture, they also enable controlled comparison between base and domain-adapted models, facilitating interpretation of the effects of biomedical fine-tuning.

#### 1.2.2 Pipeline Components and Experimental Conditions

##### Factorial Experiment

Within each LLaMa model we evaluated all combinations of:

- **Prompt strategy:** zero-shot (ZS), few-shot (FS), chain-of-thought (CoT), and extract-then-phenotype (ETP), defined below.
- **Extract-then-phenotype (ETP):** Unlike direct classification, ETP prompts the LLM to explicitly extract phenotype-relevant findings (for example, symptoms, laboratory results, and therapies) from each chunk. Extracted concepts are aggregated at the record level using phenotype-specific weights. A phenotype is deemed positive if the weighted score exceeds a fixed threshold (Figure 2). Concept lists and coefficients were determined in consultation with clinicians (details in Supplemental File 1 - Supplementary Methods).
- **Chunking:** none; naïve chunking (sentence-aware 256-token windows); and semantic chunking (section-based chunks derived via regular expressions over 12 common discharge-note headers).
- **Aggregation:** chunk-level predictions were combined using a rule based on how many chunks were classified as positive:

– any-positive (*≥*1 positive chunk)
– *≥*2-positive (at least two positive chunks)
– majority (*≥*50% of chunks positive)

**Figure 2:**
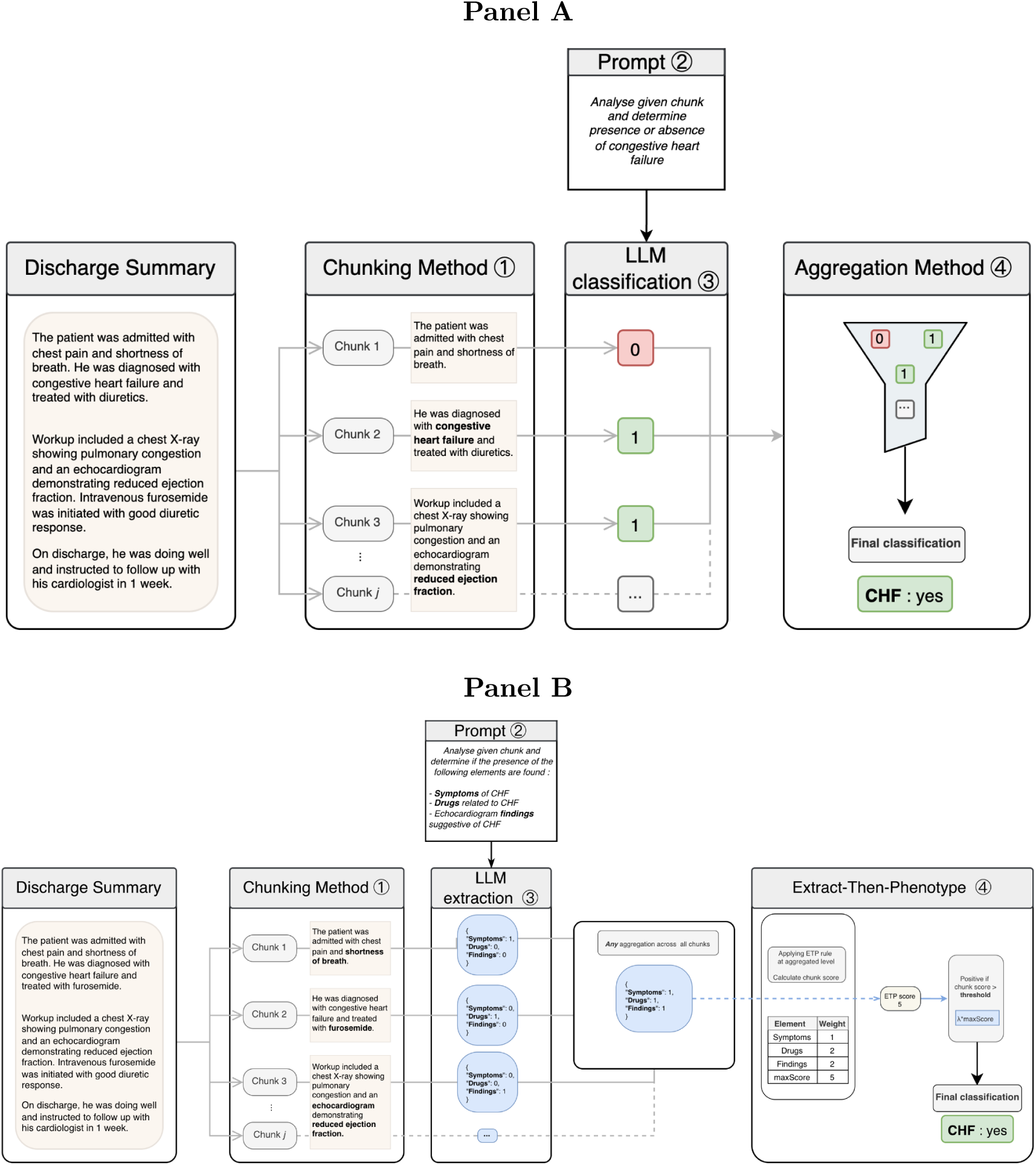
LLM-based frameworks for phenotype classification from discharge summaries. **Panel A.** A classical LLM-based classification framework. A discharge summary is segmented into multiple chunks using a selected chunking method (1). A predefined prompt (2) instructs the large language model (LLM (3)) to classify each chunk for the presence or absence of a specific phenotype, returning a binary prediction. In this example, the target phenotype is congestive heart failure (CHF). Chunk-level predictions (0 = absent, 1 = present) are then combined using an aggregation rule (4), such as majority vote. This pipeline enables systematic evaluation of how different configurations (chunking, prompts, LLMs, and aggregation methods) influence final phenotype classification accuracy. **Panel B.** The Extract-Then-Phenotype (ETP) framework. In this approach, the LLM (3) is explicitly prompted (2) to extract specific clinical elements from each chunk, such as symptoms, treatments, or diagnostic findings relevant to the target phenotype (for example, CHF). Each extracted element is assigned a predefined weight based on its diagnostic importance for the phenotype. Aggregation is then applied across all chunks. An ETP score is computed as a weighted sum of these elements (4). If the score exceeds the learned threshold (*λ ×* maxScore), the corpus is classified as positive.

The factorial experiment yields 24 configurations per model. Structural zeros arise because (i) no-chunking configurations do not require aggregation, and (ii) ETP prompting only supports its paired aggregation method. A fixed temperature of 0.5 was used for all experiments. Prompts details are provided in Supplemental File 2 - LLM Prompts and Supplemental File 3 - Extract-then-Phenotype Details.

##### Cross-Model Experiment (Fixed Configuration)

To compare model classes without pipeline confounding, we used a single configuration: CoT prompting, no chunking, no aggregation. This choice was guided by pilot evidence that this configuration provided optimal performance within the LLaMa family. DeepSeek-R1 was evaluated with zero-shot prompting as recommended for reasoning models.

### 1.3 Evaluation

The primary metric was macro-F1 across phenotypes. Secondary metrics were micro-F1, precision, recall, and accuracy. To quantify efficiency, we reported macro-F1 relative to inference cost (USD per million tokens) and an F1-efficiency metric that combines min–max–normalized macro-F1 (80% weight) with inverse cost (20% weight; formula provided in Supplemental File 1 - Supplementary Methods).

#### Statistical Analysis

We conducted a series of two-way ANOVAs on structurally complete subsets of the experiment to isolate the contribution of LLM pipeline components to phenotyping performance. We report F-statistics, Holm-adjusted p-values, and partial ² for the following anal-ysis:

1. Model *×* Chunking: assesses the effect of text segmentation strategy (no chunking, naïve chunking, semantic chunking) across models. Analysis was restricted to configurations with any-positive aggregation, and performance was aggregated across prompting strategies (zero-shot, few-shot, chain-of-thought).
2. Model *×* Aggregation: assesses the effect of aggregation rules across models. Analysis was restricted to configurations that were chunked (naïve or semantic), and performance was aggregated across prompting strategies.
3. Model *×* Prompting: assesses the effect of prompting strategy across models. Because ETP prompting strategy is only compatible with its own ETP aggregation scheme, aggregation was collapsed across {any-positive, extract-then-phenotype} to ensure a structurally valid comparison. Additional details are provided in Supplemental File 1 - Supplementary Methods.

To assess whether certain phenotypes are intrinsically harder, we also ran a two-way ANOVA (Phenotype *×* Model) and a mixed-effects model (F1 ∼ Model * Configuration + (1 | Phenotype)) treating phenotype as a random intercept.

#### Sensitivity Analysis Using Coefficient of Variation (CV)

We assessed the sensitivity of model performance to each configuration parameter (prompting, chunking, aggregation using the coefficient of variation (CV), defined as the standard deviation of macro-F1 divided by its mean. We similarly evaluated phenotype-level variability across the three LLaMA models. Higher CV values indicate greater performance variability and sensitivity to configuration choices, whereas lower CV values indicate more stable performance.

#### Pairwise Mc Nemar’s Tests

For cross-model comparisons, we used pairwise McNemar’s tests to evaluate whether classification patterns differed on the same discharge summaries across phenotypes. We applied Bonferroni correction to control the family-wise error rate across multiple comparisons.

### 1.4 Uncertainty (Entropy) Experiment

To characterize stochasticity we sampled 16 notes per phenotype (eight easy and eight challenging; criteria in Supplemental File 1 - Supplementary Methods), which yielded a total 256 observations for which we ran each factorial configuration three times and assessed Shannon entropy over predicted labels. We additionally assessed the coefficient of variation across parameters and ran one-way ANOVAs for each factor (details in Supplemental File 1 - Supplementary Methods).

#### Self-Confidence Calibration

Using a variant of the fixed CoT prompt strategy, we prompted LLaMa 3B/8B/70B/405B to qualitatively self-report confidence (low/medium/high) and evaluate calibration by stratifying F1 within confidence bins. Prompt details appear in Supplemental File 2 - LLM Prompts.

#### Error Analysis

We manually reviewed a stratified sample of misclassified cases by the largest models under the fixed configuration to develop a qualitative taxonomy of errors. We assessed the respective justifications and reasoning outputs of the model to characterize the source of classification errors.

## 1.5 Results

### 1.5.1 1. Performance Overview

We evaluated nine language models across 1,388 discharge summaries and 16 phenotypes, spanning variations in prompting strategy, chunking method, and aggregation rule. Analyses proceeded in two phases:

1. a factorial experiment within the LLaMa family (3B, 8B, 70B) to isolate configuration effects, and
2. a fixed-configuration comparison across open-source, fine-tuned, commercial, and reasoning models.

Each LLaMA model was evaluated under 24 pipeline configurations (see Figure S1), while additional models were assessed under a standardized configuration.

Table 2 summarizes macro-F1 performance for each configuration parameter (Chunking, Aggregation, Prompting) across the three LLaMA models. LLaMA models exhibited the expected scaling trend, with macro-F1 increasing from 0.53 (3B) to 0.66 (8B) and 0.86 (70B). Although absolute performance varied across phenotypes, highlighting that some phenotypes were difficult even for the largest models.

**Table 2:** Effect of Pipeline Parameters on Phenotyping Performance.

| Parameter | Level | LLaMa 3B | LLaMa 8B | LLaMa 70B | Inter-model<br>macroF1 |
| --- | --- | --- | --- | --- | --- |
| Chunking | None | 0.567 | 0.706 | 0.837 | $0.703 \pm 0.110$ |
| | Naive | 0.413 | 0.524 | 0.630 | $0.522 \pm 0.089$ |
| | Document | 0.395 | 0.492 | 0.586 | $0.491 \pm 0.078$ |
| *Intra-model macroF1 ± SD* | | $0.458 \pm 0.094$ | $0.574 \pm 0.115$ | $0.684 \pm 0.134$ | |
| Aggregation | Any | 0.525 | 0.610 | 0.727 | $0.621 \pm 0.083$ |
| | $\geq 2$ | 0.500 | 0.625 | 0.723 | $0.616 \pm 0.091$ |
| | Majority | 0.236 | 0.344 | 0.428 | $0.336 \pm 0.079$ |
| *Intra-model macroF1 ± SD* | | $0.413 \pm 0.132$ | $0.534 \pm 0.130$ | $0.639 \pm 0.142$ | |
| Prompting | Zero-Shot | 0.435 | 0.572 | 0.636 | $0.548 \pm 0.084$ |
| | Few-Shot | 0.444 | 0.506 | 0.639 | $0.530 \pm 0.081$ |
| | CoT | 0.427 | 0.536 | 0.646 | $0.536 \pm 0.089$ |
| | ETP | 0.392 | 0.556 | 0.678 | $0.542 \pm 0.117$ |
| *Intra-model macroF1 ± SD* | | $0.425 \pm 0.023$ | $0.543 \pm 0.028$ | $0.650 \pm 0.019$ | |

*Note.* Across the LLaMa family (3B–70B), two-way ANOVAs showed large and significant main effects of Model ( $\eta_p^2 = 0.53; 0.86; 0.86$ ) and both Chunking ( $\eta_p^2 = 0.97$ ) and Aggregation ( $\eta_p^2 = 0.84$ ). The effect of Prompting was smaller and not statistically significant ( $p = 0.158, \eta_p^2 = 0.185$ ). No significant Model × Parameter interactions were found for Chunking, Aggregation, or Prompting, indicating that parameter rankings are qualitatively consistent across model sizes.

For the cross-model comparison, DeepSeek-R1 achieved the highest overall macro-F1 (0.891), followed by GPT-4o and LLaMA-405B, which were statistically indistinguishable (Table 3). Ad-ditional descriptive statistics for each configuration are provided in the supplementary materials (Table S1, Table S2).

**Table 3:** Model Comparison Under a Fixed Pipeline.

| Performance Group | Models (statistically equivalent) | Macro-F1 (per model) |
| --- | --- | --- |
| Group 1 (Top Performance) | DeepSeek-R1 | 0.8914 |
| Group 2 (High Performance Cluster) | LLaMa-70B / GPT-4o / LLaMa-405B | 0.8538 / 0.8519 / 0.8517 |
| Group 3 | Gemini Flash 2.0 | 0.8331 |
| Group 4 (Mid-Tier Equivalent) | LLaMa-8B / OpenBio-70B | 0.7196 / 0.7179 |
| Group 5 | LLaMa-3B | 0.6183 |
| Group 6 (Lowest Performance) | OpenBio-8B | 0.2793 |

*Note.* Macro-F1 values are averaged over 16 phenotypes. Performance groups reflect statistically indistinguishable models under Bonferroni-corrected McNemar tests ( $\alpha < 0.05$ ). Groups are ordered by Macro-F1, and each group significantly outperforms all lower groups.

### 1.5.2 2. Effects of Pipeline Parameters on Phenotyping Performance

#### 2.1 Main Effects

Across pooled two-way ANOVAs, model size showed a large and consistent main effect on performance (²p = 0.53–0.86), with larger models achieving higher macro-F1 across all configurations (70B > 8B > 3B; Table 2).

Among pipeline components, chunking strategy had the strongest influence on performance (²p = 0.97). Evaluations performed without chunking consistently achieved the highest macro-F1, whereas both naïve and document-level chunking substantially degraded performance (none > naïve > document; Table 2). This effect was robust across model sizes.

Aggregation strategy also exerted a large effect (²p = 0.84). Aggregation rules that preserved recall outperformed majority voting, which consistently produced the lowest macro-F1. Specifically, the two-vote rule marginally outperformed the any-positive rule, while majority voting resulted in a marked drop in performance (Table 2).

In contrast, prompting strategy had the weakest impact on performance (²p = 0.19; p = 0.16). Zero-shot, few-shot, and chain-of-thought prompting performed similarly, while the extract-then-phenotype approach underperformed all direct classification strategies.

### 1.5.3 2.2 Interactions Effects and Robustness Across Model Sizes

Across all two-way ANOVAs, no Model *×* Parameter interactions were statistically significant, indicating that the relative ordering of chunking, aggregation, and prompting strategies was stable across model sizes. Although larger models achieved higher absolute performance, pipeline design choices affected small and large models in the same direction. Accordingly, the highest-performing configuration across all LLaMA models was whole-document inference without chunking, combined with any aggregation and either zero-shot or chain-of-thought prompting.

These findings were supported by the coefficient-of-variation (CV) analysis (Figure S1). Aggregation method accounted for the largest relative variability in performance (26.3%), followed by phenotype (23.3%), whereas prompting contributed the least variability (approximately 4–6%), mirroring its weak main effect in the ANOVA. CV analysis further showed that the 70B model exhibited uniformly lower variability across all parameters, indicating greater robustness to pipeline design choices compared with smaller models.

### 1.5.4 3. Phenotype-Level Effects on Performance

Phenotypes varied widely in baseline difficulty, with mean macro-F1 across models ranging from approximately 0.40 for the most challenging conditions (e.g., C. difficile complications) to 0.90 for the easiest (e.g., hypertension). A two-way Phenotype *×* Model ANOVA confirmed a strong main effect of phenotype (F15,1089 = 23.1, p < 10^-54^), demonstrating substantial intrinsic variability in how difficult the 16 phenotypes are to identify from discharge summaries. Detailed phenotype-level performance are provided in the Supplementary Material (Figure S2–S4).

Despite this broad variation in absolute difficulty, the relative effect of model size was consistent across all phenotypes. Larger models provided uniform performance gains (70B *≥* 8B *≥* 3B), with no Phenotype *×* Model interaction detected (p = 0.91). A mixed-effects model treating phenotype as a random intercept reinforced this result (ICC 0.38), indicating that configuration effects were invariant to phenotype. In other words, the optimal configuration—zero-shot prompting, no chunking, and any-aggregation—was the same for all phenotypes.

Phenotype-wise CV analysis complemented these findings (Figure 3). Some phenotypes, including deep vein thrombosis and pulmonary embolism complications (DVT/PE) and C. difficile complications, exhibited high sensitivity to model capacity (CV > 30%), whereas others, such as hypertension and rheumatoid arthritis, showed low variability across models (CV < 10%). This pattern indicates that configuration choice and model scale have disproportionately large effects on difficult phenotypes, even though the optimal configuration and model ranking remain constant.

**Figure 3:**
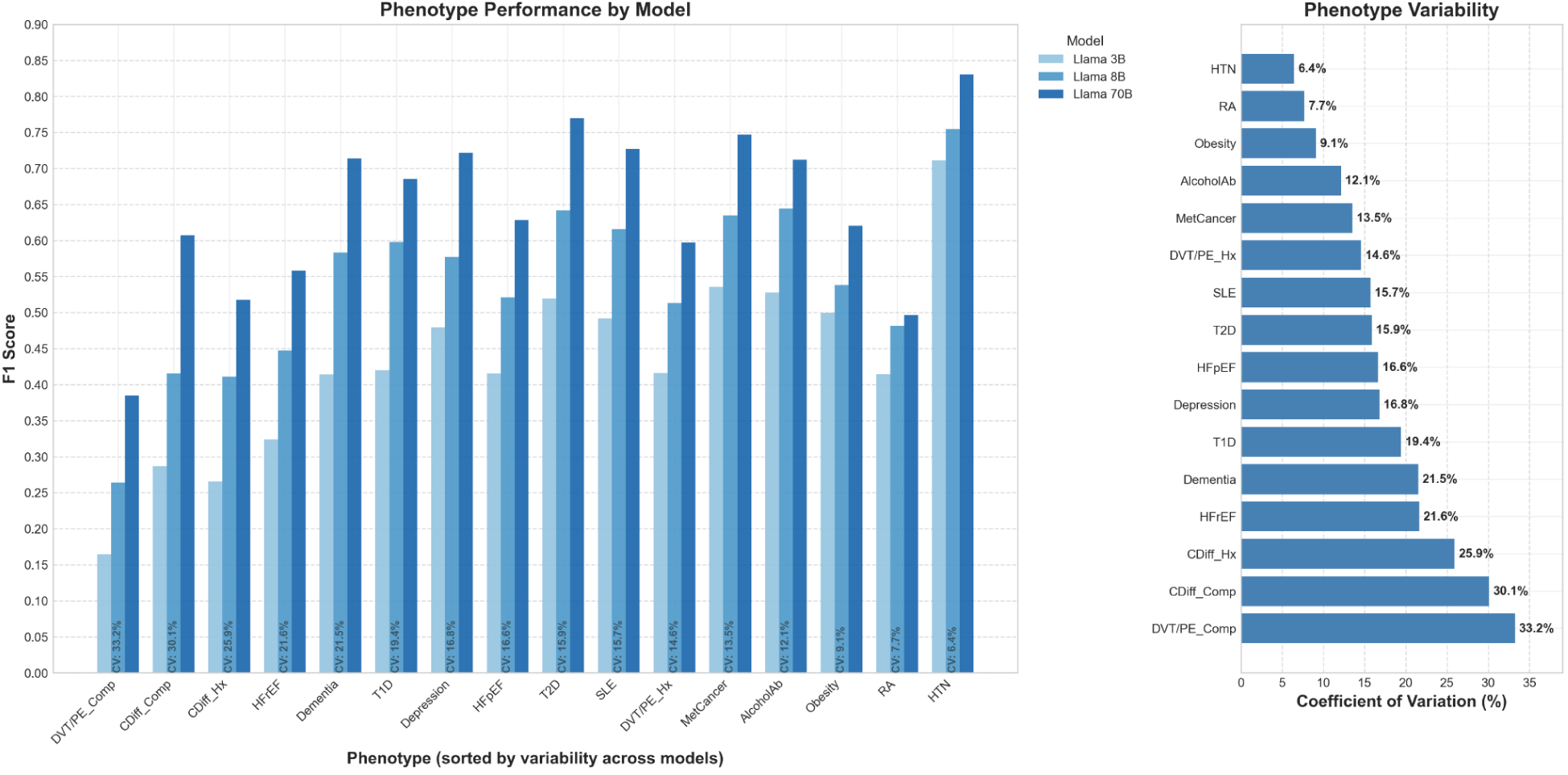
Phenotype-level variation in model performance and sensitivity. *Note.* The right panel shows the coefficient of variation (CV) across model capacities and configurations. Low-variability phenotypes (CV < 10%) correspond to easier phenotyping tasks with consistently high F1 scores, where even smaller models perform well (for example, RA and HTN). High-variability phenotypes (CV > 25%) represent harder tasks, with lower baseline performance and greater sensitivity to model choice (for example, DVT/PE complications). The left panel illustrates that increasing model size yields minimal gains for easier phenotypes but substantial improvements for harder ones. Overall, phenotype difficulty is intrinsic: model rankings remain stable, but the benefit of additional capacity grows with task difficulty.

Taken together, these results show that phenotype difficulty is intrinsic, and while larger models yield consistent improvements across all phenotypes, the magnitude of benefit is greatest for the most challenging clinical conditions.

### 1.5.5 4. Cross-Model Comparison Under a Fixed Configuration

To isolate intrinsic model capabilities, we evaluated all models under a standardized pipeline: chain-of-thought (CoT) prompting with whole-document inference, without chunking or aggregation (zero-shot prompting for DeepSeek-R1, as recommended for reasoning-oriented models). Pairwise McNemar’s tests with Bonferroni correction (36 comparisons; = 0.001389) identified six statistically distinct performance groups (Table 3).

DeepSeek-R1 formed the top-performing group, significantly outperforming all other models. LLaMA-70B, GPT-4o, and LLaMA-405B constituted a statistically indistinguishable high-performance cluster. Gemini Flash 2.0 achieved the strongest mid-tier performance, followed by a lower mid-tier cluster (LLaMA-8B and OpenBioLLM-70B), while LLaMA-3B and OpenBioLLM-8B formed the lowest-performing group.

These results confirm that under equivalent inference conditions, model scale and architecture exert the dominant influence on phenotyping accuracy

### 1.5.6 5. Additional Analyses

#### 5.1 Prediction Entropy and Uncertainty

Prediction entropy analyses (Figure S5) identified the primary determinants of model uncertainty. One-way ANOVAs revealed strong effects of task difficulty, chunking strategy, model size, prompting, and aggregation (all p < 0.05), with phenotype difficulty explaining the largest share of variance. In contrast, sampling temperature had a statistically detectable but practically negligible effect (p = 0.047; <0.01 entropy units).

Larger models consistently produced lower entropy, indicating more stable predictions, and whole-document inference yielded lower entropy than chunked configurations—mirroring its superior macro-F1 performance. In absolute terms, entropy values were low across all conditions, with all mean entropy values < 0.3.

Coefficient-of-variation (CV) analysis reinforced these findings. Phenotype difficulty (CV 94%), model size (62%), and chunking method (61%) accounted for most uncertainty variability, whereas prompting and aggregation contributed minimally. Together, these results show that prediction uncertainty, like accuracy, is primarily shaped by model capacity and input structure, rather than prompt design.

#### 5.2 Self-Confidence Calibration

Self-reported confidence was moderately informative across all models (Figure 4). Predictions labeled as high confidence consistently achieved the highest empirical performance (F1 0.82–0.88), whereas medium-confidence predictions were poorly calibrated.

**Figure 4a:**
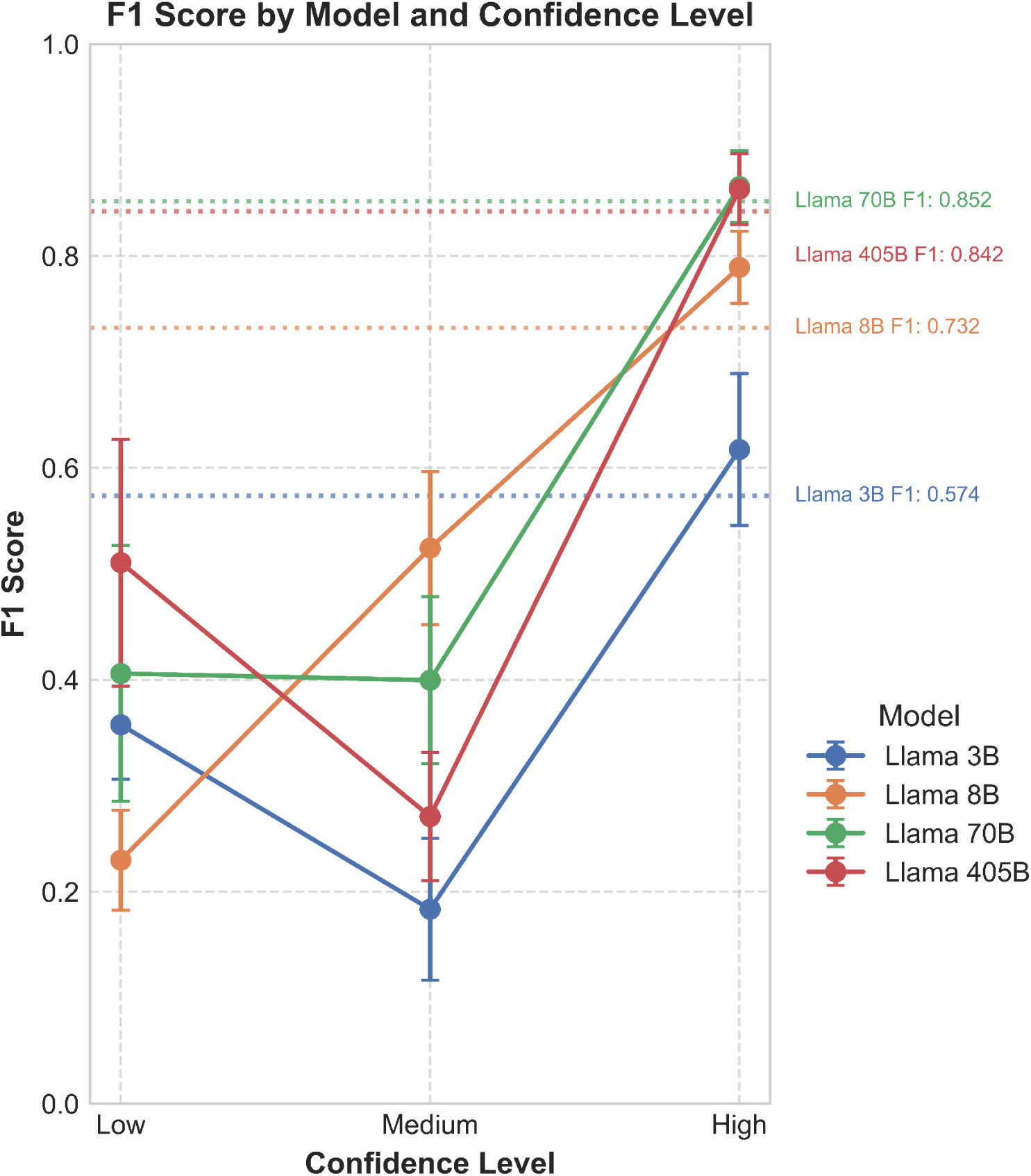
Self-confidence calibration vs. F1 Across Models. *Note.* This figure shows how model-reported confidence (low, medium, high) relates to actual F1 performance across four LLaMA models. Error bars show standard deviation across phenotypes. Dashed lines indicate each model’s overall mean F1. F1 rises sharply with confidence for all models, with LLaMA-70B and LLaMA-405B exceeding 0.85 F1 when predictions are labeled high-confidence. Medium-confidence outputs are unreliable across models, and low-confidence outputs perform poorly.

**Figure 4b:**
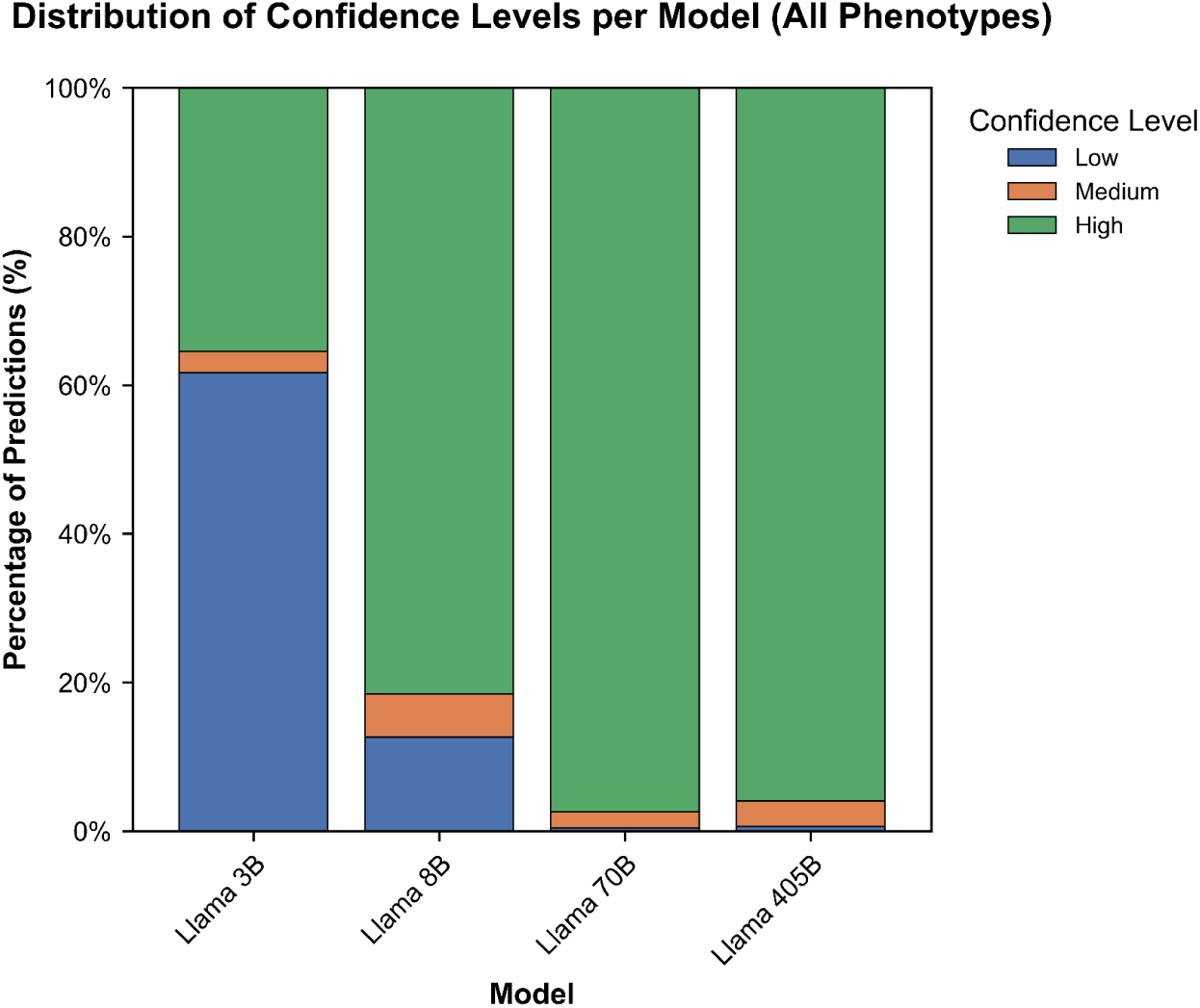
Distribution of Confidence Levels Across Models. *Note.* This stacked bar plot displays the proportion of predictions each model labeled as low, medium, or high confidence. Larger models (LLaMA-70B, LLaMA-405B) classify over 95% of predictions as high-confidence, whereas LLaMA-3B assigns low confidence in most cases. These patterns highlight how confidence calibration improves with model scale and how smaller models express greater uncertainty in their predictions, but this is overshadowed by a tendency for overconfidence in larger models. Smaller models (LLaMA-3B and LLaMA-8B) exhibited the greatest miscalibration, frequently assigning high confidence to incorrect predictions. In contrast, larger models, including DeepSeek-R1, LLaMA-70B, and GPT-4o, demonstrated a more reliable monotonic relationship between confidence and accuracy. However, systematic overconfidence persisted across all models, with large models assigning high confidence to the majority of outputs.

#### 5.3 Cost–performance Trade-Offs

Inference cost varied substantially across models. The cost-performance trade-off illustrates a nonlinear tradeoff, where performance gains beyond mid-tier open models require disproportionate cost increases (see Figure S6). DeepSeek-R1 achieved the highest macro-F1, while maintaining moderate inference cost relative to large commercial models. GPT-4o delivered strong performance but at the highest per-token cost, resulting in comparatively poor cost-efficiency.

In contrast, LLaMA-70B and LLaMA-405B achieved competitive performance at substantially lower cost, placing them in the most favorable region of the performance–cost Pareto frontier. Fine-tuned biomedical models (OpenBioLLM-8B and OpenBioLLM-70B) did not provide a performance advantage and incurred higher cost per unit of accuracy, rendering them dominated options in this comparison.

### 1.5.7 6. Error Analysis

To characterize failure modes in LLM-based phenotyping, we conducted a targeted manual review of 57 misclassified observations using cases correctly classified by DeepSeek-R1 while all other models failed, enabling isolation of reasoning-specific advantages; and cases misclassified by all models, including DeepSeek-R1, capturing instances of universal model failure. For each case, we manually reviewed the full discharge summary, model predictions, and reasoning traces (see Supplemental File 3 - Extract-then-Phenotype Details).

This analysis revealed a small number of recurring error mechanisms that accounted for most failures across phenotypes:

- **Temporal disambiguation failures**, where historical conditions were misinterpreted as active or salient negative tests overrode prior positive evidence.
- **Ontological granularity errors**, in which related but distinct entities (e.g., cutaneous lupus vs. systemic lupus) were conflated.
- **Inference gaps**, where models failed to compute derived quantities (e.g., BMI) or interpret laboratory thresholds in the absence of explicit diagnostic terms.
- **Diagnostic certainty mismatches**, where tentative or hedged statements were treated as confirmed diagnoses contrary to ground-truth criteria.
- **Abbreviation misinterpretation**, reflecting incorrect expansion or contextual understanding of clinical acronyms.

We should note that we did not encounter discharge summary structural issues as a source of error. Table 4 summarizes this error taxonomy, with additional details provided in Supplemental File 4 - Error Analysis.

**Table 4:** Taxonomy of Phenotyping Errors Observed in LLM-Based EHR Phenotyping.

| Error Category | Mechanism of Failure | Primary Source of Error | Representative Example |
| --- | --- | --- | --- |
| Temporal disambiguation failure | LLM fails to distinguish historical from active conditions; overly weights recent negative evidence | Model reasoning (temporal logic) | Phenotype: C. difficile (history)<br>Text: "History of C. Diff colitis." + "Current PCR negative."<br>Model: Predicts negative by letting current test override historical evidence. |
| Ontological granularity error | LLM treats a related subtype as equivalent to the target phenotype | Prompt or guidance | Phenotype: Systemic lupus erythematosus (SLE)<br>Text: "Subcutaneous (cutaneous) lupus."<br>Model: Predicts SLE, failing to distinguish cutaneous vs systemic disease. |
| Inference gap | Model fails to derive a phenotype from numerical data (vitals, labs) when no explicit diagnostic term is present. | Model does not infer phenotype based on indirect evidence | Phenotype: Obesity<br>Text: "Height 5'3", Weight 102.7 kg (BMI 40)."<br>Model: Predicts negative because BMI was not computed; no "obese" token present. |
| Diagnostic Certainty Threshold | LLM interprets tentative/hedged language as positive evidence, but ground truth requires confirmed diagnosis | Model interpretation | Phenotype: Metastatic cancer<br>Text: "Lesions suspicious for metastasis."<br>Model: Predicts positive; ground truth requires confirmed metastasis. |
| Abbreviation misinterpretation | LLM applies literal acronym expansion, ignoring medication patterns or demographic context | Model misunderstanding of clinical usage patterns | Phenotype: Diabetes type 2<br>Text: "History of IDDM." + "Medications: Metformin, Glipizide."<br>Model: Predicts Type 1 (IDDM), but context strongly suggests insulin dependent diabetes in patients with type 2 diabetes. |

## 1.6 Discussion

In this study, we systematically evaluated how pipeline design choices shape large language model (LLM) performance for EHR phenotyping. Using a factorial experimental design, we identify which parameters materially influence accuracy and reliability—and which do not. Three principal findings emerge.

First, input structure is the primary determinant of phenotyping performance. Across all models, configurations using whole-document inference consistently outperformed both naïve and document-based chunking strategies. This is because discharge summaries encode interdependent clinical information across multiple sections; fragmenting this context disrupts cross-sectional dependencies and increases both local false negatives and false positives^22^. Among chunked configurations, recall-preserving aggregation rules (any or two or more aggregation) outperformed majority voting, which suppressed true signals confined to individual chunks. In contrast, prompting strategy contributed little to performance, with zero-shot, few-shot, and chain-of-thought prompting yielding similar results across models and phenotypes. These findings indicate that prompt engineering does not compensate for poor segmentation and that preserving global clinical context is critical for accurate phenotyping.

Second, model capacity and architectural design exert a strong and consistent influence on performance. Within the LLaMA family, phenotyping accuracy scales predictably with model size, with diminishing returns beyond approximately 70B parameters^23^. Cross-model comparisons conducted under a fixed configuration further showed that DeepSeek-R1 outperformed larger frontier models, while LLaMA-70B achieved performance comparable to GPT-4o and LLaMA-405B at substantially lower cost. This pattern suggests that architectural features—particularly those supporting reason-ing—can meaningfully affect phenotyping accuracy once scale saturates^24,25^. These results do not imply that any single architecture is universally optimal, but underscore that model design matters beyond parameter count alone. Those findings are compatible with similar growing evidence of the limits of the scaling law in LLMs^24^. Additionally, biomedical domain-adapted models offered worse performance than their non fine-tuned counterparts in our study, a surprising finding that was also previously documented in the literature suggesting that poor training data negatively affect performance of these models on a wide variety of tasks (clinical reasoning, summarization, entity extraction).^26^

Third, phenotype difficulty in intrinsic, yet optimal pipeline configurations are remarkably stable across clinical conditions. Absolute performance varied widely across phenotypes, reflecting differences in temporal complexity, clinical reasoning requirements, and documentation clarity. However, neither model ranking nor the highest-performing configuration changed by phenotype. Mixed-effects modeling confirmed that while phenotype-level heterogeneity accounted for substantial variance, configuration effects were invariant across phenotypes. Phenotype-wise variability analyses further showed that the most challenging phenotypes benefited disproportionately from increased model capacity and higher-performing configurations.

We found no relationship between phenotype prevalence and macro-F1, nor between phenotype type (clinical vs social determinant of health), limiting generalizability of simple heuristics for defining phenotype difficulty. Instead, some differentiators were (a) phenotypes requiring inference (i.e. HFrEF requires numerical interpretation of echocardiogram report), (b) phenotypes require temporal reasoning (C. diff and DVT) and those with (c) relative conceptual ambiguity (i.e. obesity).

We further evaluated aspects relevant to deployment, including output stability under repeated sampling, model-reported confidence and cost. Prediction entropy was low overall and driven primarily by phenotype difficulty rather than prompt choice or sampling temperature. Larger models and whole-document configurations produced lower entropy, mirroring their superior performance. Together, these results showcase the reliability of LLMs for large-scale classification tasks.

Model-reported confidence was only moderately informative: high-confidence predictions were more accurate on average, but larger models frequently assigned high confidence to incorrect predictions. This systematic overconfidence indicates that confidence should not be interpreted as calibrated probability; instead, confidence is best used to support triage and auditing (e.g., routing low-confidence or discordant cases to manual review) rather than as a stand-alone acceptance criterion.

Cost–performance analyses provide additional operational context. While DeepSeek-R1 achieved the highest macro-F1, LLaMA-70B and LLaMA-405B occupied the most favorable region of the performance–cost Pareto frontier^27^. These results emphasize that model selection should consider both cost alongside accuracy, especially in a large scale deployment scenario

Error analysis identified recurring mechanisms underlying model failures across phenotypes. These included temporal disambiguation errors, ontological conflation of related entities, numeric inference gaps, diagnostic certainty mismatches, and abbreviation misinterpretation. Importantly, even the highest-performing models exhibited these errors, demonstrating that strong aggregate performance does not eliminate clinically meaningful failure modes. This taxonomy supports practical safeguards: targeted audits for high-risk phenotypes, explicit logic constraints for known temporal and numeric pitfalls and explicit definition of thresholds for phenotyping classifications.

These findings translate into concrete deployment implications for data warehouse phenotyping pipelines (Box 1).

### Box 1. Practical implications for deploying LLM phenotyping pipelines

#### Recommended defaults

**Prefer whole-document inference** when the note fits within the model context window; treat chunking as a deliberate design choice rather than a default.

**Use a single, standardized prompt template across phenotypes** (e.g., ZS or CoT with structured output). This improves reproducibility in data warehouse pipelines.

**Select models using a performance–cost criterion** (macro-F1 vs inference cost). Reserve larger models for intrinsically difficult phenotypes and note that fine-tuned biomedical models are not inherently better for this task.

Align deployment mode with governance: self-hosted inference is generally preferred for PHI-bearing notes; API-based inference requires explicit institutional approvals and controls.

#### When to deviate

**Chunking:** If operational constraints require chunking (e.g., context limits/longitudinal EHR notes use), **avoid majority voting** and prefer recall-preserving aggregation (e.g., any-positive; consider *≥*2-positive when false positives have high downstream cost).

Consider retrieval techniques to select relevant passages, although this would require appropriate internal validation to ensure adequate performance.

**Prompt strategy:** Maintain a common baseline prompt, but introduce **minimal phenotype-specific constraints** when motivated by observed failure modes—particularly for phenotypes requiring (i) **temporal logic** (history vs active), (ii) **numeric inference** (e.g., BMI/EF thresholds), or (iii) **abbreviation/ontology disambiguation**.

Any phenotype-specific prompt modifications should be validated and compared against the baseline approach.

##### Confidence handling and decision policy

**Do not treat self-reported confidence as calibrated probability.** Use confidence primarily for **triage and auditing** (e.g., prioritize low-confidence cases for review), not as a stand-alone acceptance criterion.

**Adopt a tiered decision policy:**

**Auto-accept** predictions only in settings with strong phenotype-specific validation and low harm from errors;

**Route to human review** for difficult phenotypes, borderline evidence, or discordant signals (e.g., hedged language, conflicting temporal cues, numeric thresholds).

**Continuously monitor known failure modes** in production audits: temporal disambiguation, diagnostic certainty thresholds, numeric inference gaps, ontological granularity errors, and abbreviation misinterpretation.

#### Validation expectations

**Validate at the phenotype level** (not only macro-averaged performance), and stratify evaluation by difficulty where possible. Performance gains from larger models are often greatest on the hardest phenotypes, but these are also the phenotypes where error costs may be highest.

This study has limitations. We restricted inputs to a single discharge summary per admission, which limits direct generalization to longitudinal, multi-note EHR phenotyping pipelines^28^. That said, our findings align with Wornow et al. in that input selection/structure (what text the model sees) often matters more than elaborate prompting for cohort-identification tasks.

However, this design isolates the contribution of pipeline components independently of retrieval and note selection. While effect magnitudes may differ in multi-note settings, the central principle—that increased context discontinuity degrades recall—is likely to generalize. Additional limitations include the single-institution dataset and English-only clinical notes. We also recognize that a proper phenotype taxonomy for classifying is needed to guide researchers to preemptively estimate the inherent difficulty of a given phenotype, which we were not powered to do in this study.

In summary, this work provides a systematic characterization of LLM pipeline design for EHR phenotyping. Across diverse phenotypes, performance is governed primarily by input structure and model capacity rather than prompt engineering, and the highest-performing configurations are stable across clinical conditions. Together with uncertainty, cost, and error analyses, these results provide evidence-based guidance for building reproducible phenotyping pipelines while underscoring the need for phenotype-level validation and human oversight despite relatively high performance of LLM for phenotyping tasks.

## Supporting information

S3 Extract-then-Phenotype Approach Details

S4 Error Analysis

S2 LLM Prompts

## 1.7 Data Availability

The MIMIC-IV Phenotype Atlas (MIPA) dataset labels are openly available at: https://github.com/open-health-data-lab/MIPA-datacard

All code for running the LLM experiment phenotyping is available at https://github.com/ open-health-data-lab/MIPA

Note that access to the underlying discharge summaries and clinical notes is conditional upon completing CITI training and obtaining authorized access to the MIMIC-IV v2.2 database through PhysioNet.

## 1.8 Author Contributions

Eric Yamga: Conceptualization, methodology, data analysis, and manuscript drafting.

Sean Murphy: Conceptualization, manuscript editing.

Philippe Després: Conceptualization, critical review, and manuscript editing.

## Data Availability

The MIMIC-IV Phenotype Atlas (MIPA) dataset labels are openly available at https://github.com/open-health-data-lab/MIPA-datacard
All code for pipeline processing, supervised model training, and LLM-based phenotyping is available at https://github.com/open-health-data-lab/MIPA
Note that access to the underlying discharge summaries and clinical notes is conditional upon completing CITI training and obtaining authorized access to the MIMIC-IV v2.2 database through PhysioNet.

