## Supplementary material for "A Systematic Exploration of LLM Behavior for EHR Phenotyping": S1 Supplementary Methods

### 1 Supplemental File 1 - Supplementary Methods

**A) LLM pipeline components** This section details the technical specifications of the large-language-model (LLM) phenotyping pipeline and the parameters varied in the factorial experiment. The pipeline comprises three components: prompting strategy, chunking strategy, and aggregation strategy.

**A.1 Models, access method, and inference settings** All LLM experiments were executed using the OpenRouter API, which provides a unified interface to multiple commercial and open-source LLM providers while preserving provider-specific model identifiers. For reproducibility, we report the exact OpenRouter model strings used at inference time. The only exception was for the biomedical models, for which runs were done via the HuggingFace API. All runs were executed October–November 2025.

##### Frontier / Commercial Models

- **GPT-4o:** `openai/gpt-4o-2024-11-20`
- **Gemini Flash 2.0:** `google/gemini-2.0-flash-001`

##### Open-Source LLaMA Family (Instruction-Tuned)

- **LLaMA-3B:** `meta-llama/llama-3.2-3b-instruct`
- **LLaMA-8B:** `meta-llama/llama-3.1-8b-instruct`
- **LLaMA-70B:** `meta-llama/llama-3.3-70b-instruct`
- **LLaMA-405B:** `meta-llama/llama-3.1-405b-instruct`

##### Reasoning Model

- **DeepSeek-R1 (full):** `deepseek/deepseek-r1`

##### Biomedical Domain-Adapted Models

- **OpenBioLLM-8B:** `aaditya/Llama3-OpenBioLLM-8B`
- **OpenBioLLM-70B:** `aaditya/Llama3-OpenBioLLM-70B`

##### Inference Parameters

- **temperature** = 0.5
- **max_tokens** = 3000
- **top_p** = provider default (not overridden)

No random seed control was available through the API for inference.

**Retry / JSON Validity Policy:** Each inference call was attempted up to 3 times when the output failed JSON parsing. After each failure, we applied a JSON-repair routine and retried. If parsing still failed after 3 attempts, the run was excluded from analysis.

**A.2 Chunking strategy** We evaluated three strategies for segmenting discharge summaries before inference:

**No Chunking (Whole-Document Inference):** The full discharge summary was passed to the model as a single input. All MIPA discharge summaries fit within model context limits; no truncation was required. No aggregation step applies.

**Naïve Token-Bounded Chunking:** Notes were split into non-overlapping windows capped at 256 model tokens. Sentence boundaries were respected to avoid splitting mid-sentence.

**Semantic Header-Aware Chunking (Document):** Notes were segmented into clinically meaningful sections using case-insensitive regular expressions over 12 common discharge-note headers (optional trailing colon and whitespace). This chunking mode is referred to as "document" in tables/figures:

- History of Present Illness
- Past Medical History
- Family History
- Social History
- Allergies
- Admission Physical Exam
- Discharge Physical Exam
- Impression
- Brief Hospital Course
- Discharge Condition
- Discharge Instructions
- Discharge Medications

**A.3 Prompting strategies** We evaluated four prompting strategies:

**Zero-Shot (ZS):** Direct binary decision prompt (Yes/No) without exemplars.

**Few-Shot (FS):** ZS template preceded by three curated exemplars.

**Chain-Of-Thought (Co T):** Prompt instructing the model to provide brief clinical justification (e.g., using symptoms, medications, labs, prior diagnoses) and then output a binary decision.

**Extract-Then-Phenotype (ETP):** A structured approach where the LLM extracts phenotype-relevant findings from each chunk (based on phenotype-specific curated concept lists), concepts are unioned across chunks, and a deterministic record-level decision rule is applied using phenotype-specific weights.

**ETP Decision Rule (a Priori Threshold):** A record is classified as positive if the weighted sum of extracted concepts exceeds a fixed threshold defined a priori: threshold = 0.4 × (maximum possible score). Concept lists and concept weights were defined a priori by consensus among two clinician–informaticians.

**A.4 Aggregation strategy (map–reduce pipelines)** For chunked pipelines using ZS/FS/-CoT, chunk-level binary predictions were combined into a record-level label using one of the following rules:

| Method | Description |
|---|---|
| Any-positive | Record positive if ≥1 chunk is positive |
| At-least-two | Record positive if ≥2 chunks are positive |
| Majority vote | Record positive if >50% of chunks are positive |
| ETP record-level scoring | Specific to the ETP framework.<br>For each chunk, the LLM extracts phenotype-relevant elements (concepts) from a phenotype-specific list. Extracted concepts are unioned across all chunks to form a single record-level set (any-aggregation), which is then converted into an ETP score by summing predefined concept weights.<br><br>The record is classified as positive if the ETP score exceeds a fixed threshold: 0.4 × maxScore (the maximum possible score for that phenotype). This approach aggregates evidence across chunks at the concept level, not by voting over chunk-level labels. |

**B) Statistical analysis** All analyses were conducted in Python 3.11 using statsmodels and pingouin.

**B.1 Configuration effects (factorial experiment)** The factorial experiment varied model size, prompting, chunking, and aggregation. Some combinations are structurally invalid (e.g., no chunking implies no aggregation; ETP prompting is only defined with ETP scoring). To avoid structurally impossible cells while preserving interpretability, we performed two-way ANOVAs on structurally valid subsets using Type-II sums of squares with HC3 robust standard errors.

**(1) Model × Chunking** Assesses the effect of segmentation (none, naïve, semantic/document) across models. Analysis was restricted to any-positive aggregation, and performance was aggregated across ZS/FS/CoT prompting.

**(2) Model × Aggregation** Assesses the effect of aggregation rules across models. Analysis was restricted to chunked configurations (naïve or semantic/document), and performance was aggregated across ZS/FS/CoT prompting.

**(3) Model × Prompting** Prompting strategies are not all compatible with the same aggregation schemes: ETP prompting is only defined with ETP record-level scoring, whereas ZS/FS/CoT operate with map–reduce aggregation rules (any-positive, at-least-two, majority). As a result, a full Model × Prompt comparison across all aggregation rules would contain structurally impossible cells.

To enable a Model × Prompt analysis on a structurally valid subset, we constructed the comparison as follows:

- **Prompt levels included:** {ZS, FS, CoT, ETP}.
- **Map–reduce aggregation:** Represented by any-positive (used for ZS/FS/CoT).
- **ETP record-level scoring:** Used for ETP.
- **Operational implementation:** Aggregation was grouped as {any-positive, ETP}, ensuring that each prompt strategy was evaluated with its valid aggregation mechanism.
- **Chunking handling:** Chunking was allowed to vary across its levels to avoid conditioning results on a single segmentation strategy.

This design supports estimation of the main effects of Model and Prompt, and the Model × Prompt interaction, which tests whether specific prompting strategies differentially benefit (or impair) particular model families or sizes.

We report F-statistics, Holm-adjusted p-values, and partial $\eta^2$.

**B.2 Phenotype-Level Effects** To assess heterogeneity across clinical conditions, we performed:

- A two-way ANOVA (Phenotype × Model) on macro-F1.
- A mixed-effects model with phenotype treated as a random intercept to quantify phenotype-level heterogeneity (ICC reported in the main Results). A full Phenotype × Model × Configuration ANOVA was not estimable due to rank deficiency.

**B.3 Coefficient of Variation (CV)** To quantify relative sensitivity of macro-F1 to design choices, we computed the coefficient of variation:

$$\mathrm{CV} = \left( \frac{\mathrm{SD}(F1)}{\mathrm{Mean}(F1)} \right) \times 100\%$$

**B.4 F1-Efficiency Metric** To balance performance and cost, we defined an efficiency score:

$$\text{Normalized F1} = \frac{F1_i - F1_{\text{min}}}{F1_{\text{max}} - F1_{\text{min}}}$$

$$\text{Normalized Cost} = 1 - \frac{\text{Cost}_i - \text{Cost}_{\text{min}}}{\text{Cost}_{\text{max}} - \text{Cost}_{\text{min}}}$$

$$\text{Efficiency} = 0.8 \times \text{Normalized F1} + 0.2 \times \text{Normalized Cost}$$

**C) Entropy Analysis** We conducted a complementary analysis to quantify stochasticity and consistency of LLM predictions under repeated sampling. We used Shannon entropy computed from repeated outputs:

$$H(X) = -\sum P(x) \log_2 P(x)$$

Lower entropy indicates more consistent predictions across repeated runs.

**C.1 Case selection (easy vs challenging)** For each phenotype, we selected 16 discharge summaries: 8 easy and 8 challenging, based on precomputed case difficulty labels derived from external model agreement and configuration-level stability.

**Easy Cases Were Defined As** Discharge summaries where the phenotype was correctly classified:

- by both GPT-4o and LLaMA-405B, and
- by the other benchmark models (LLaMA-3B/8B/70B, Gemini Flash 2.0) in more than 70% of their configuration runs.

**Challenging Cases Were Defined As** Admissions where the phenotype was misclassified:

- by both GPT-4o and LLaMA-405B, and
- by the other benchmark models (LLaMA-3B/8B/70B, Gemini Flash 2.0) in more than 50% of their configuration runs.

**C.2 Experimental design and factors** Each (note, phenotype, configuration) combination was evaluated with 3 repeated inference runs to estimate variability. Entropy experiments were run for LLaMA-3B, LLaMA-8B, and LLaMA-70B, and at temperatures $\{0.0, 0.5, 1.0\}$.

**Entropy Was Analyzed as a Function of**

- Temperature (0.0, 0.5, 1.0)
- Model size (3B, 8B, 70B)
- Task difficulty (easy vs challenging)
- Chunking method (none, naïve, semantic/document)
- Prompt strategy (ZS, FS, CoT, ETP)
- Aggregation rule (any-positive, at-least-two, majority, ETP scoring)

We used one-way ANOVAs to quantify the effect of each factor on entropy (Type-II sums of squares; =0.05; Holm correction for multiple testing).

**D) Error Analysis** To characterize failure modes, we performed manual review under a fixed cross-model configuration:

- CoT prompting + no chunking for all models that produced justification strings.
- Zero-shot prompting for DeepSeek-R1, with reasoning tokens retained in outputs.

**D.1 Sampling Strategy** We sampled two strata of cases, each randomly stratified across phenotypes:

- **Tier A (n=30):** cases where DeepSeek-R1 was correct and other models failed (reasoning advantage cases).
- **Tier B (n=30):** cases where all models, including DeepSeek-R1, disagreed with the reference label (universal failure cases).

**D.2 Review procedure** A single reviewer examined the discharge summary, model prediction, and justification/reasoning traces. Errors were coded into a taxonomy of recurrent mechanisms (reported in Table 4), with full case narratives provided in Supplementary Material 2.
