## Supplementary material for "A Systematic Exploration of LLM Behavior for EHR Phenotyping": S3 Extract-then-Phenotype Approach Details

### 1 Supplemental File 3 - Extract-then-Phenotype Details

C Diff (Past)

| Element | Element Category | Description | Individual JSON Key | Weight |
|---|---|---|---|---|
| Explicit mention of CDI | Diagnosis | Documented diagnosis of Clostridium difficile infection. | c_diff | 2 |
| Recurrence | Diagnosis | Documentation of recurrent or multiple past episodes | recurrent | 2 |
| Prior treatment | Treatment | Previous treatment with oral vancomycin, fidaxomicin, or metronidazole | prior_treatment | 1 |
| Fecal transplant | Treatment | Use of fecal microbiota transplant as a treatment option. | prior_fmt | 1 |
| Past complication from CDI | Treatment | Past complications from CDI (e.g., toxic megacolon, colectomy) | prior_complications | 1 |

C Diff (Complication)

| Element | Element Category | Description | Individual JSON Key | Weight |
|---|---|---|---|---|
| New CDI Diagnosis | Diagnosis | Explicit mention of new diagnosis of C. difficile, C. diff, CDI, or pseudomembranous colitis during admission | phenotype | 2 |
| CDI Symptoms | Clinical | New onset severe diarrhea, abdominal pain/cramping, fever | symptoms | 1 |
| Diagnostic Testing | Diagnostic | Positive C. difficile toxin test, PCR, or GDH assay during admission | diagnostic_tests | 2 |
| Colonoscopy Findings | Diagnostic | Pseudomembranes or other CDI findings on colonoscopy during admission | colonoscopy | 1 |

| Element | Element Category | Description | Individual JSON Key | Weight |
|---|---|---|---|---|
| CDI specific treatment | Treatment | Use of oral vancomycin for CDI treatment or fidaxomicin or metronidazole | treatment | 2 |
| CDI Complications | Complications | Development of complications (toxic megacolon, shock, colectomy) | complications | 1 |
| FMT Required | Treatment | Requirement for fecal microbiota transplantation | fmt_needed | 1 |
| Nosocomial Infection | Classification | Clear evidence infection developed >48 hours after admission | nosocomial | 1 |

#### Diabetes II

| Element | Element Category | Description | Individual JSON Key | Weight |
|---|---|---|---|---|
| Explicit mention of Type 2 Diabetes | Diagnosis | Documented diagnosis of Type 2 Diabetes. | diabetes_type_2 | 2 |
| Use of oral hypoglycemic agents | Medication | Use of medications such as metformin, sulfonylureas, DPP-4 inhibitors, GLP-1 agonists. | metformin, sulfonylureas, dpp4_inhibitors, glp1_agonists, sglt2_inhibitors | 2 |
| Insulin use | Medication | Use of insulin in management, especially in advanced stages of Type 2 Diabetes. | insulin | 1 |
| Diabetic complications | Complications | Presence of complications such as neuropathy, nephropathy, or retinopathy. | neuropathy, nephropathy, retinopathy | 1 |
| HbA1c > 6.5% | Laboratory Results | HbA1c levels greater than 6.5%, indicative of diabetes. | hba1c | 1 |
| Blood Glucose Level (Random) | Laboratory Results | Venous or capillary glucose> 200 mg/dL | glucose_levels | 1 |

#### Diabetes I

| Element | Element Category | Description | Individual JSON Key | Weight |
|---|---|---|---|---|
| Explicit mention of Type I Diabetes | Diagnosis | Documented diagnosis of Type I Diabetes. | diabetes_type_1 | 2 |
| Insulin therapy from diagnosis | Medication | Dependence on insulin from the time of diagnosis, typical for Type I diabetes. | insulin therapy from diagnosis | 2 |
| Autoantibodies | Laboratory Results | Presence of specific anti-islets antibodies such as anti-GAD, anti-ZnT8, anti-IA2, and anti-Insulin antibodies, which support a diagnosis of Type I diabetes. | autoantibodies | 1 |
| Ketoacidosis | Complications | Occurrence of diabetic ketoacidosis, a severe complication often seen in Type I diabetes. | ketoacidosis | 2 |
| Insulin therapy | Medication | Any use of insulin | insuline therapy any | 1 |
| No oral hypoglycemics | Medication | Absence of oral hypoglycemic agents | no oral hypoglycemics | 2 |
| Neuropathy | Complications | Presence of complications such as neuropathy. | neuropathy | 1 |
| Retinopathy | Complications | Presence of complications such as retinopathy. | retinopathy | 1 |
| Nephropathy | Complications | Presence of complications such as nephropathy. | nephropathy | 1 |

Dementia

| Element | Element Category | Description | Individual JSON Key | Weight |
|---|---|---|---|---|
| Explicit mention of Dementia | Diagnosis | Dementia (including variants like vascular dementia, AD, PD, etc.) excluding MCI | dementia | 2 |
| Cognitive decline | Symptom | Evidence of progressive cognitive decline, a core symptom of dementia. | cognitive decline | 1 |

| Element | Element Category | Description | Individual JSON Key | Weight |
|---|---|---|---|---|
| Memory loss | Symptom | Frequent or severe memory loss, especially short-term memory loss. | memory loss | 1 |
| Disorientation | Symptom | Disorientation to time and place, common in dementia. | disorientation | 1 |
| Behavioral changes | Symptom | Changes in behavior or personality, typical in dementia. | behavioral changes | 1 |
| Cholinesterase inhibitors | Medication | Use of medications like donepezil, rivastigmine, which are commonly prescribed for dementia. | cholinesterase inhibitors | 2 |
| Memantine | Medication | Use of memantine, specifically for moderate to severe cases. | memantine | 2 |
| Neuroimaging findings | Diagnostic Indicator | MRI or CT findings that support a diagnosis of dementia, such as atrophy in specific brain areas. | neuroimaging findings | 1 |
| Functional decline | Symptom | Loss of ability to perform daily activities independently, indicating progression of dementia. | functional decline | 2 |

Depression

| Element | Element Category | Description | Individual JSON Key | Weight |
|---|---|---|---|---|
| Explicit mention of Depression | Diagnosis | Documented diagnosis of depression. | depression | 2 |
| Mood symptoms | Symptom | Sadness, emptiness, anhedonia, loss of interest/pleasure. | mood symptoms | 1 |
| Sleep disturbances | Symptom | Changes in sleeping patterns, either insomnia or hypersomnia, associated with depression. | sleep disturbances | 1 |

| Element | Element Category | Description | Individual JSON Key | Weight |
|---|---|---|---|---|
| Appetite changes | Symptom | Significant weight loss or gain due to changes in appetite, common in depression. | appetite changes | 1 |
| Antidepressants | Medication | Use of specific medications, including SSRIs, SNRIs, TCAs, MAOIs, or other antidepressants. | antidepressants | 2 |
| Psychotherapy | Treatment | Engagement in psychotherapy sessions as part of treatment. | psychotherapy | 1 |
| Fatigue | Symptom | Reports of significant fatigue or loss of energy nearly every day. | fatigue | 1 |
| Suicide risk | Symptom | Suicidal thoughts, plans, or attempts. | suicide risk | 1 |

HTN

| Element | Element Category | Description | Individual JSON Key | Weight |
|---|---|---|---|---|
| Explicit mention of Hypertension | Diagnosis | Documented diagnosis of hypertension or high blood pressure. | hypertension | 2 |
| Blood pressure readings | Diagnostic Indicator | Documented high blood pressure readings (e.g., systolic $\geq$130 mmHg or diastolic $\geq$80 mmHg), or description of blood pressure as ”elevated” or ”high”. | blood pressure readings | 2 |
| Antihypertensive medications | Medication | Use of specific medications, including ACE inhibitors, ARBs, beta-blockers, calcium channel blockers, thiazide diuretics, and other antihypertensives. | antihypertensive medications | 2 |

| Element | Element Category | Description | Individual JSON Key | Weight |
|---|---|---|---|---|
| End organ damage caused by hypertension | Complications | • Renal: Impaired kidney function<br>• Ocular: Hypertensive retinopathy<br>• Cardiac: Left ventricular hypertrophy, heart failure<br>• Vascular: Coronary artery disease, peripheral artery disease<br>• Cerebral: Stroke, transient ischemic attack (TIA) | end-organ_damage | 1 |

HfpEF

| Element | Element Category | Description | Individual JSON Key | Weight |
|---|---|---|---|---|
| Explicit mention of Heart Failure with Preserved Ejection Fraction | Diagnosis | Documented diagnosis of Heart Failure with Preserved Ejection Fraction. | hfpef | 2 |
| Preserved EF | Diagnostic Indicator | EF >50% or described as preserved/normal. | preserved_EF | 2 |
| Diastolic dysfunction | Diagnostic Indicator | Mentioned in echocardiography or other cardiac imaging. | diastolic_dysfunction | 2 |
| Congestion symptoms | Symptom | Dyspnea, edema, orthopnea, fatigue. | congestion_symptoms | 1 |
| Elevated filling pressures | Diagnostic Indicator | E/e' ratio >14, elevated LAP or LVEDP. | elevated_filling_pressures | 1 |
| NT-proBNP levels | Diagnostic Indicator | NT-proBNP levels (> 900 pg/mL) or BNP levels (> 100 pg/mL) | bnp_levels | 1 |
| Use of diuretics | Treatment | Use of diuretics to manage fluid retention specifically furosemide (Lasix) in the context of HFpEF. | diuretics | 1 |

HFrEF

| Element | Element Category | Description | Individual JSON Key | Weight |
|---|---|---|---|---|
| Explicit mention of Heart Failure with Reduced Ejection Fraction | Diagnosis | Documented diagnosis of Heart Failure with Reduced Ejection Fraction. | hfref | 2 |
| Reduced EF | Diagnostic Indicator | EF <40% or described as reduced/low. | reduced_EF | 2 |
| Congestion symptoms | Symptom | Dyspnea, edema, orthopnea, fatigue. | congestion_symp-toms | 1 |
| NT-proBNP levels | Diagnostic Indicator | NT-proBNP levels (> 900 pg/mL) or BNP levels (> 100 pg/mL) | bnp_levels | 1 |
| Use of beta-blockers | Treatment | Prescription of beta-blockers to improve survival in patients with HFrEF. | beta_blockers | 1 |
| Use of ARNI | Treatment | Use of angiotensin receptor-neprilysin inhibitors to enhance cardiac function and reduce heart failure hospitalizations. | arni | 1 |
| Use of diuretics | Treatment | Use of diuretics to manage fluid retention specifically furosemide (Lasix) in the context of HFrEF. | diuretics | 1 |
| Use of ACE inhibitors or ARBs | Treatment | Prescription of ACE inhibitors or ARBs to improve outcomes in patients with HFrEF. | acei_arb | 1 |
| Use of aldosterone antagonists | Treatment | Prescription of aldosterone antagonists to improve outcomes in patients with HFrEF. | aldosterone_antag-onists | 1 |

#### Lupus

| Element | Element Category | Description | Individual JSON Key | Weight |
|---|---|---|---|---|
| Explicit mention of SLE | Diagnosis | Documented diagnosis of SLE, lupus, or systemic lupus erythematosus. | sle | 2 |
| Positive ANA test | Laboratory Results | Positive antinuclear antibodies test (titer < 1:180). | ANA | 2 |
| Specific antibodies | Laboratory Results | Positive Anti-dsDNA, anti-Sm, anti-phospholipid antibodies. | specific_antibodies | 2 |
| Low complement levels | Laboratory Results | Low C3 or C4 complement levels. | complement_levels | 1 |
| Skin manifestations | Symptoms | Specific cutaneous manifestations of systemic lupus: malar rash, discoid rash, photosensitivity. | skin_manifestations | 1 |
| Joint symptoms | Symptoms | Arthritis, arthralgia in multiple joints. | joint_symptoms | 1 |
| Renal involvement | Complications | Explicit mention of lupus nephritis or proteinuria, hematuria, indications of lupus nephritis Class II, III, IV, or V. | renal_involvement | 1 |
| Serositis | Complications | Pleuritis, pericarditis. | serositis | 1 |
| Use of DMARDs | Medication | hydroxychloroquine, methotrexate, mycophenolate mofetil, azathioprine, belimumab, or cyclophosphamide. | dmards | 2 |
| Use of corticosteroids | Medication | Use of corticosteroids | steroids | 1 |

#### Metastatic cancer

| Element | Element Category | Description | Individual JSON Key | Weight |
|---|---|---|---|---|
| Explicit mention of metastatic cancer | Diagnosis | Documented diagnosis of solid metastatic cancer, stage IV cancer, or advanced cancer. | metastatic_cancer | 2 |

| Element | Element Category | Description | Individual JSON Key | Weight |
|---|---|---|---|---|
| Primary tumor site | Diagnosis | Identification of the primary tumor location. | primary tumor site | 1 |
| Metastasis sites | Diagnosis | Documented locations of metastasis, such as bones, liver, lungs, or brain. | metastasis sites | 2 |
| Biopsy results | Diagnostic Results | Histological confirmation from biopsy of metastatic sites. | biopsy results | 1 |
| Oncological treatments | Treatment | Use of treatments specific to metastatic cancer, such as chemotherapy, targeted therapy, or immunotherapy. | oncological treatments | 1 |

RA

| Element | Element Category | Description | Individual JSON Key | Weight |
|---|---|---|---|---|
| Explicit mention of rheumatoid arthritis | Diagnosis | Documented diagnosis of rheumatoid arthritis, RA, or terminology indicating chronic inflammatory joint disease. | rheumatoid_arthritis | 2 |
| Joint symptoms | Symptoms | Swelling, pain, stiffness in multiple joints, especially hands and feet. | joint_symptoms | 1 |
| Morning stiffness | Symptoms | Stiffness lasting >1 hour in the morning. | morning_stiffness | 1 |
| Serological markers | Laboratory Results | Rheumatoid factor (RF), anti-cyclic citrullinated peptide (anti-CCP) antibodies. | serological_markers | 2 |
| Imaging findings | Diagnostic Results | Joint erosions, narrowing of joint spaces on X-rays, MRI, or ultrasound. | imaging_findings | 1 |

| Element | Element Category | Description | Individual JSON Key | Weight |
|---|---|---|---|---|
| cDMARDs | Treatment | Use of conventional disease-modifying antirheumatic drugs (e.g., methotrexate, sulfasalazine, leflunomide). | cdmards | 2 |
| Biologics | Treatment | Use of biologic agents (e.g., TNF inhibitors, rituximab, abatacept, tocilizumab). | biologics | 2 |
| Steroids | Treatment | Use of corticosteroids for RA management (e.g., prednisone, methylprednisolone). | steroids | 1 |
| Extra-articular manifestations | Complications | Rheumatoid nodules, lung involvement, vasculitis. | extra-articular_manifestations | 1 |

Obesity

| Element | Element Category | Description | Individual JSON Key | Weight |
|---|---|---|---|---|
| Explicit mention of obesity | Diagnosis | Documented diagnosis of obesity or terminology indicating excessive body weight. | obesity | 2 |
| BMI | Diagnostic Results | Documented Body Mass Index (BMI) categorizing the patient as obese (BMI 30 or higher) or inferred based on weight and height available | BMI | 2 |
| Waist circumference | Diagnostic Results | Measurement of waist circumference indicating abdominal obesity (greater than 40 inches in men and 35 inches in women). | waist_circumference | 1 |

| Element | Element Category | Description | Individual JSON Key | Weight |
|---|---|---|---|---|
| Comorbid conditions | Complications | Presence of obesity-related comorbidities such as type 2 diabetes, hypertension, or dyslipidemia, OSA | comorbid_conditions | 1 |
| Bariatric surgery | Treatment | History of or planned bariatric surgery for weight loss. | bariatric_surgery | 2 |
| Medication for obesity | Treatment | Use of medications specifically aimed at weight loss (e.g., orlistat, liraglutide, ozempic). | medication_for_obesity | 1 |

ROH Abuse

| Element | Element Category | Description | Individual JSON Key | Weight |
|---|---|---|---|---|
| Explicit mention of alcohol abuse | Diagnosis | Documented diagnosis of alcohol abuse, alcohol use disorder, or alcoholism. | alcohol_abuse | 2 |
| Consumption patterns | Symptoms | Heavy or frequent alcohol use, binge drinking. | consumption_patterns | 1 |
| Withdrawal symptoms | Symptoms | Signs of alcohol withdrawal such as tremors, agitation, or seizures. | withdrawal_symptoms | 1 |
| Liver abnormalities | Diagnostic Results | Abnormal liver enzymes, signs of alcoholic liver disease (labs or imaging). | liver_abnormalities | 1 |
| Comorbid conditions | Complications | Alcohol-related pancreatitis, gastritis, alcoholic liver disease. | comorbid_conditions | 2 |
| Treatment for addiction | Treatment | Involvement in alcohol addiction treatment programs or counseling. | treatment_for_addiction | 1 |
| Medications | Treatment | Use of disulfiram, naltrexone, acamprosate. | medications | 1 |

VTE (Past)

| Element | Element Category | Description | Individual JSON Key | Weight |
|---|---|---|---|---|
| Explicit mention of past VTE | Diagnosis | Explicit mention of any VTE (DVT, PE, SVT, CVST) | phenotype | 2 |
| Past DVT | Diagnosis | Prior deep vein thrombosis | dvt_past | 0 |
| Past PE | Diagnosis | Prior pulmonary embolism | pe_past | 0 |
| Past SVT | Diagnosis | Prior splanchnic venous thrombosis (portal, mesenteric, splenic, hepatic veins) | svt_past | 0 |
| Past CVST | Diagnosis | Prior cerebral venous sinus thrombosis | cvst_past | 0 |
| Recurrent VTE | Diagnosis | Documentation of recurrent or multiple past episodes | recurrent | 2 |
| Prior Treatment | Treatment | Previous or active anticoagulation treatment for past history of VTE (heparin, low molecular weight heparin, warfarin, DOACs (e.g., apixaban, rivaroxaban) | anticoagulation | 1 |
| IVC Filter History | Intervention | History of IVC filter placement | ivc_filter | 1 |
| Thrombophilia | Risk Factor | Known thrombophilia or hypercoagulable condition | thrombophilia | 1 |

VTE (Complication)

| Element | Element Category | Description | Individual JSON Key | Weight |
|---|---|---|---|---|
| Explicit mention of New VTE | Diagnosis | New diagnosis of any VTE during admission (DVT, PE, SVT, CVST) | phenotype | 2 |
| New DVT | Diagnosis | New deep vein thrombosis during admission | dvt_current | 0 |
| New PE | Diagnosis | New pulmonary embolism during admission | pe_current | 0 |
| New SVT | Diagnosis | New splanchnic venous thrombosis during admission | svt_current | 0 |

| Element | Element Category | Description | Individual JSON Key | Weight |
|---|---|---|---|---|
| New CVST | Diagnosis | New cerebral venous sinus thrombosis during admission | cvst_current | 0 |
| VTE Symptoms | Clinical | Clinical presentation (leg swelling, chest pain, abdominal pain, headache) | vte_symptoms | 1 |
| Diagnostic Testing | Diagnostic | Positive imaging or elevated D-dimer | diagnostic_tests | 2 |
| Current Treatment | Treatment | Anticoagulation initiated for new VTE during admission | anticoagulation | 2 |
| VTE Interventions | Intervention | Specific interventions (thrombolysis, thrombectomy, IVC filter) | interventions | 1 |
| Nosocomial VTE | Classification | Clear evidence VTE developed >48 hours after admission | nosocomial | 1 |
