## Supplementary material for "A Systematic Exploration of LLM Behavior for EHR Phenotyping": S4 Error Analysis

### 1 Supplemental File 4 - Error Analysis

#### Comprehensive Error Analysis

**Sampling Strategy for Error Case Review** To investigate the mechanisms driving model errors, we performed a structured manual review of discrepant predictions. Because the full experiment generated more than 22,000 phenotype-level predictions across nine models, we used a targeted sampling strategy designed to identify the most informative excerpts.

We evaluated predictions from four high-performing general-purpose models (LLaMA-70B, GPT-4o, LLaMA-405B, and Gemini Flash 2.0) and compared them against a reasoning-optimized model (DeepSeek-R1). Based on concordance and discordance of prediction errors, misclassified cases were partitioned into two analytically distinct tiers.

**Tier A: Model-Specific Reasoning Advantage** Cases where DeepSeek-R1 produced the correct phenotype label (true positive or true negative), while all other models misclassified the same case. This tier isolates scenarios where reasoning-oriented architectures outperform conventional LLMs and reveals which linguistic or clinical structures require deeper inferential processing.

- A total of 96 such cases were identified across the evaluation corpus. From these, a stratified sample of 28 cases was selected for detailed qualitative error analysis.

**Tier B: Universal Failure Cases** Cases where all models, including DeepSeek-R1, disagreed with the reference label. This tier uncovers either (a) fundamental limits of LLM comprehension or (b) dataset issues.

- A total of 284 cases met these criteria, from which a stratified sample of 29 cases was selected for further examination.

**Manual Review Protocol** Each document underwent a three-step clinician-guided audit:

- **Clinical Validity Assessment:** Reviewers read the full discharge summary, determined the clinically correct phenotype status, and compared it to the reference label.
- **Reasoning Trace Analysis (DeepSeek-R1):** For Tier A cases, the model's internal chain-of-thought was examined to evaluate whether correct predictions resulted from valid reasoning or coincidental pattern matching.
- **Cross-Model Justification Comparison:** The explanations generated by other models were compared to identify distractors, annotation-sensitive phrases, or contextual ambiguities that contributed to misclassification.

This protocol was used to classify errors and identify a taxonomy of phenotyping errors through which LLMs succeed or fail.

**Taxonomy of Phenotyping Errors** The combined Tier A and Tier B review yielded a five-category taxonomy capturing the dominant mechanisms of error (Table 4 in the main manuscript). Below, we provide expanded definitions, mechanisms, and representative examples for each class.

**S3.1 Temporal Disambiguation Failure** **Definition:** Incorrect mapping of diagnoses to the required timeframe (past versus current).

**Mechanism:**

- Misinterpretation of "history of…"
- Overweighting current laboratory results.
- Failure to apply inclusive OR logic in phenotype definitions.

**Example:** A case with "History of C. diff colitis" but a negative PCR during the admission resulted in contradictory model predictions. DeepSeek-R1 succeeded in most cases but occasionally failed because of recency bias.

**S3.2 Ontological Granularity Mismatch** **Definition:** The model identifies a subtype or related disease that is taxonomically distinct from, but linguistically overlapping with, the target phenotype.

**Mechanism:**

- Failure to interpret exclusionary modifiers such as "cutaneous" and "subcutaneous".
- Over-reliance on anchor terms such as "SLE" and "Plaquenil".

**Example:** Cases describing subcutaneous (cutaneous) lupus, correctly excluding systemic involvement, were misclassified by all models as systemic lupus erythematosus (SLE).

**S3.3 Inference Gap** **Definition:** Failure to infer diagnoses that require numerical computation, derived measurements, or diagnoses that rely on inference without an explicit diagnostic label.

**Mechanism:**

- Diagnoses based on thresholds such as BMI, hemoglobin, or creatinine/eGFR go undetected.

**Example:** A patient with height 5'3" and weight 102.7 kg (BMI $\approx$ 40) was labeled obese by ground truth, but all models, including DeepSeek-R1, predicted 0 because the word "obese" was absent.

**S3.4 Diagnostic Certainty Threshold** **Definition:** The LLM interprets tentative or hedged language as positive evidence, whereas the ground truth requires a confirmed diagnosis.

**Example:** The text mentions lesions suspicious for metastasis, but the model predicts a positive label even though the phenotype definition requires confirmed metastasis.

**S3.5 Abbreviation Misinterpretation** **Definition:** The LLM applies literal acronym expansion, ignoring medication patterns or demographic context.

**Mechanism:**

- Model misunderstanding of clinical usage patterns.

**Example:** The model predicts type 1 diabetes by interpreting "IDDM" literally, but the surrounding context strongly suggests insulin-requiring type 2 diabetes rather than autoimmune type 1 diabetes.
