## Supplementary Figures for "A Systematic Exploration of LLM Behavior for EHR Phenotyping"

### 1 Supplementary Figures

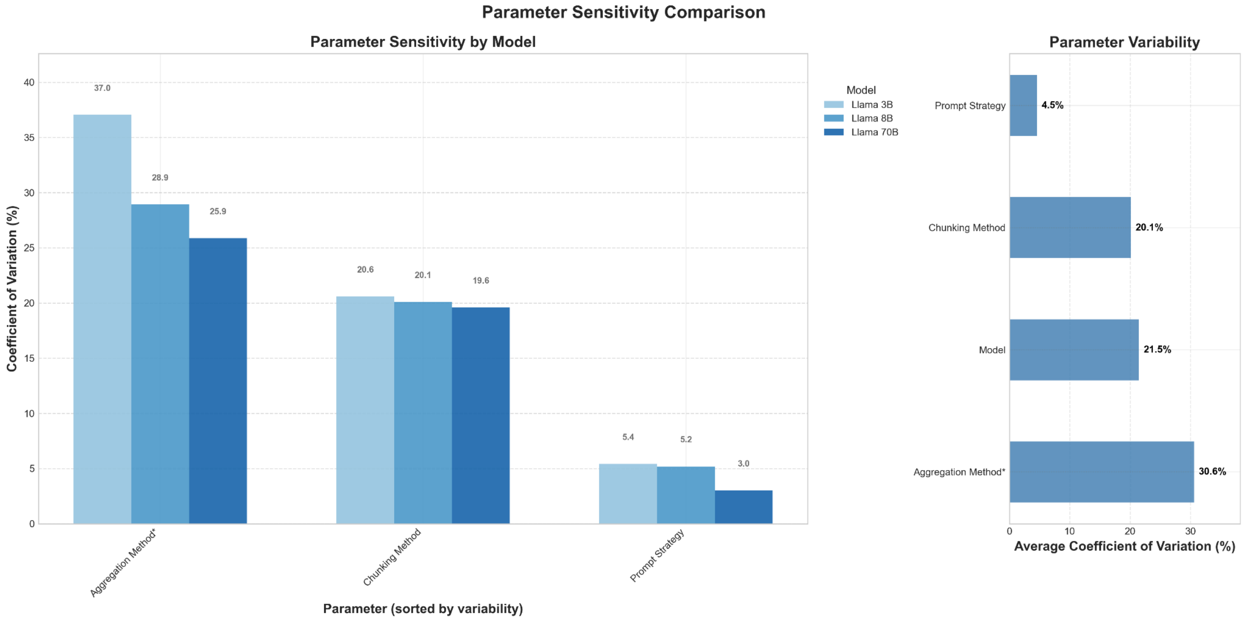


**Figure S1:** Parameter sensitivity and variability analysis using Coefficient of Variation (CV).

CV quantifies the relative variability in F1 scores as a percentage of the mean performance — for example, a CV of 4.5% for prompt strategy indicates that changing prompting methods (e.g., from zero-shot to few-shot or chain-of-thought) typically shifts F1 scores by about ±4.5% around the average value, across evaluated tasks.

The right panel presents the average CV across all models, confirming that aggregation method (30.6%), model choice (21.5%), and chunking strategy (20.1%) contribute the most to the variability in macro-F1 performance.

The left panel reports CV values for each configuration parameter stratified by model size (LLaMA 3B, 8B, 70B). Aggregation method, model choice and chunking show the highest variability, whereas prompting strategy exhibits the lowest. Consistent with the lack of significant Model × Parameter interactions, all models were impacted similarly by parameters, but effect sizes were different. Smaller models (3B, 8B) are more sensitive to changes in prompt or chunking strategy, while LLaMA-70B shows uniformly low CVs—indicating greater robustness to pipeline design choices.

*Note.* Aggregation CV excludes runs with chunking = none, and ETP prompting is omitted because it pairs exclusively with its own aggregation rule.

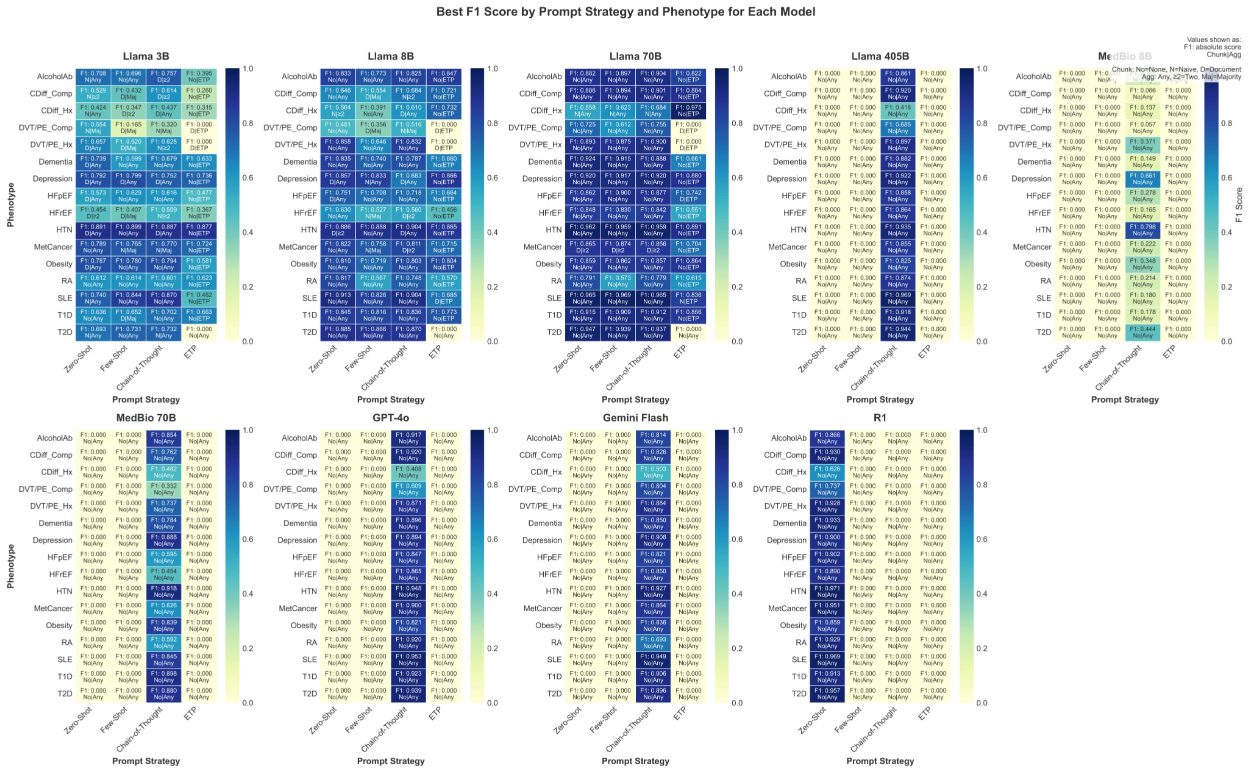


**Figure S2:** Heatmap showing best F1 scores by prompt strategy and phenotype for each model, with associated chunking and aggregation method.

This heatmap displays the highest F1 score achieved by each model–phenotype pair, stratified by prompting strategy (Zero-Shot, Few-Shot, Chain-of-Thought, Explain-then-Predict). Each cell includes the F1 score and the best-performing configuration, reported as Chunking|Aggregation. The consistent pattern of high scores under No Chunking (No) and Any Aggregation (Any) across models—especially LLaMA 70B, GPT-4o, and Gemini Flash—suggests these settings optimize classification across diverse clinical phenotypes. ETP prompting is less frequently associated with optimal performance, while Chain-of-Thought and Zero-Shot strategies appear more robust across phenotypes. Several phenotypes (e.g., T2D, HTN) achieve >0.90 F1 with large models, while others (e.g., RA, SLE) remain more challenging. Missing values (F1 = 0) indicate that no successful prediction was generated under tested configurations.

#### Legend

- **Prompt Strategy:** Zero-Shot, Few-Shot, Chain-of-Thought (CoT), Explain-then-Predict (ETP)
- **Chunking:** No = None, N = Naive, D = Document
- **Aggregation:** Any, 2 = Voting ($\geq 2$), Maj = Majority Vote

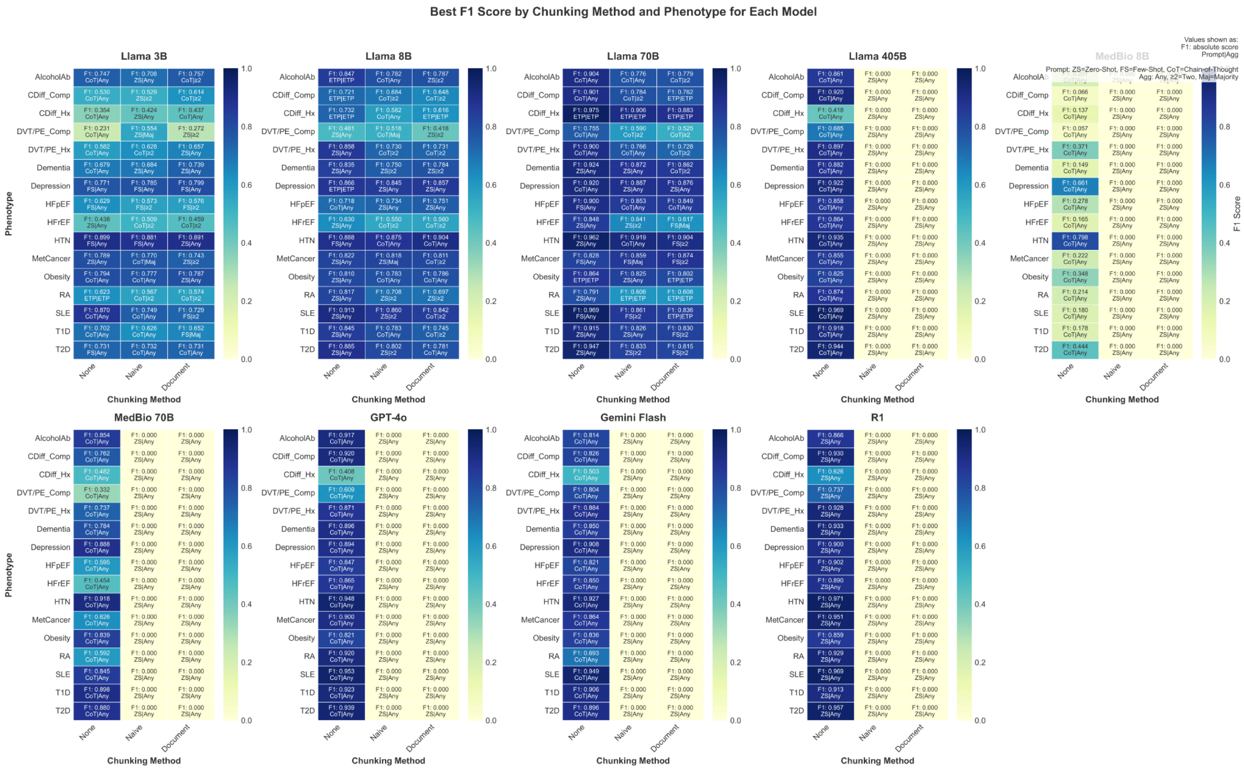


**Figure S3:** Heatmap showing best F1 Score by Chunking Method and Phenotype for Each Model

This heatmap displays the highest F1 score achieved for each combination of clinical phenotype (y-axis) and chunking method (x-axis), stratified across different language models (panels). Each cell represents the best-performing configuration under a specific chunking method, annotated with the F1 score and the associated prompt and aggregation strategy (Prompt|Agg). Prompt strategies include Zero-Shot (ZS), Few-Shot (FS), Chain-of-Thought (CoT), and Explanation-to-Prediction (ETP); aggregation strategies include Any, 2 (exactly two agreeing outputs), and Maj (majority voting).

The plot reveals significant performance variability across models and chunking approaches, with more advanced models (e.g., GPT-4o, MedBio 70B) consistently achieving higher scores. Notably, the effectiveness of chunking strategies is phenotype-dependent: chunking improves performance for some phenotypes (e.g., DVT/PE_Comp, T2D) but offers minimal or even negative returns for others (e.g., CDiff_Hx, RA).

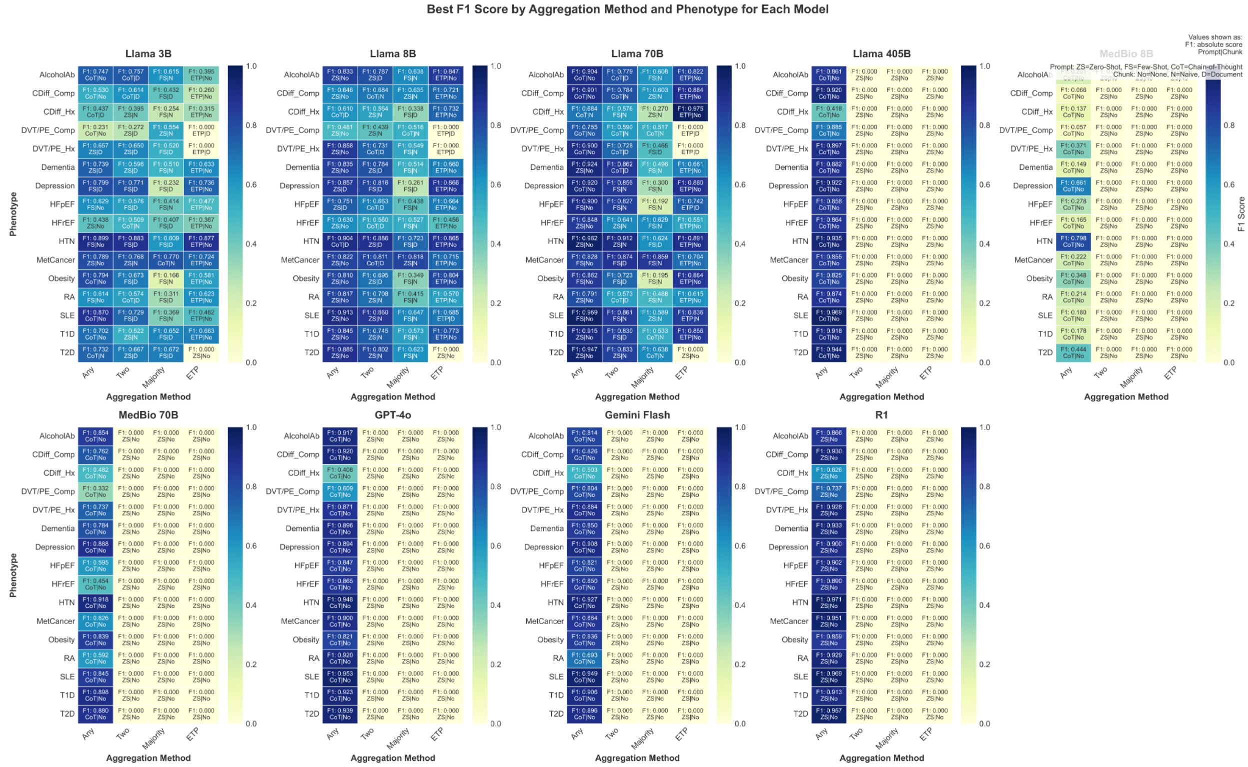


**Figure S4:** Heatmap showing best F1 score per phenotype and model across aggregation methods.

This heatmap illustrates the best-performing aggregation method for each phenotype across language models, showing the F1 score alongside the corresponding prompting and chunking strategy (e.g., CoT|D = Chain-of-Thought prompt with Document-level chunking). Aggregation methods compared include ”Any Two,” ”Majority,” and ”ETP” (Evidence-Triage Protocol). While larger models (e.g., LLaMa-70B, GPT-4o) often benefit from Chain-of-Thought prompting with no chunking and ETP-based aggregation, smaller models show heterogeneous preferences. Performance dropouts (F1 = 0) in some cells indicate either task failure or ineffective aggregation under certain configurations. This figure underscores the importance of model-specific aggregation design to optimize performance on phenotype classification tasks.

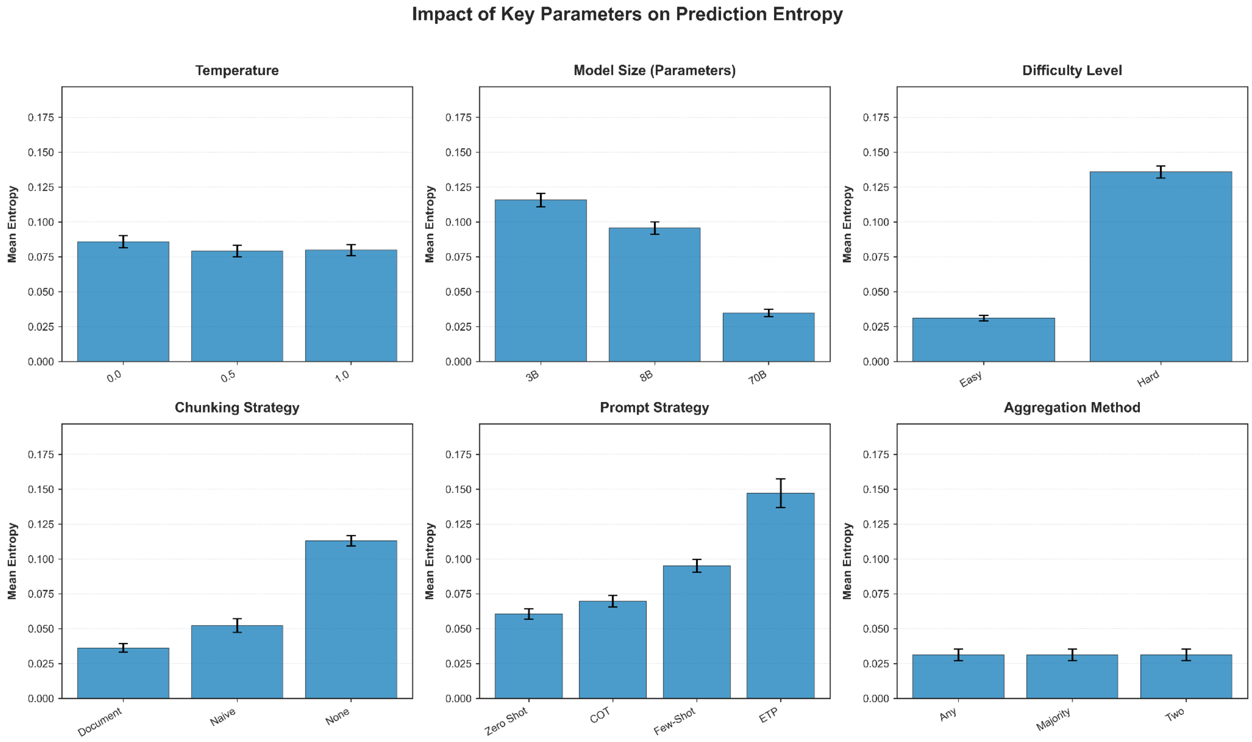


**Figure S5a:** Impact of Pipeline Parameters on Prediction Entropy

This figure shows the mean entropy of model predictions across key parameter dimensions, with error bars representing the standard error of the mean (SEM). Lower entropy indicates more consistent predictions. Results are based on an experiment in which each configuration was run three times on a 240-sample subset, varying one parameter at a time while holding all others constant.

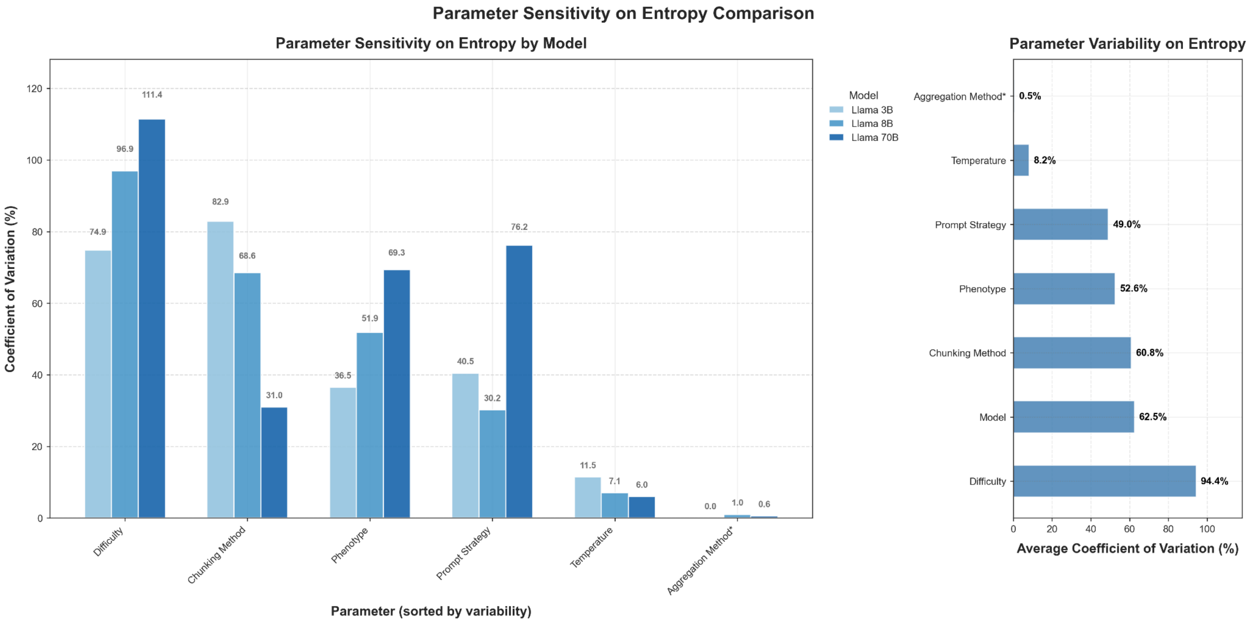


**Figure S5b:** Parameter Sensitivity in Entropy (Coefficient of Variation Analysis)

This figure summarizes how prediction entropy varies with pipeline parameters, quantified using the coefficient of variation (CV). Difficulty is the dominant source of entropy variability (CV 94.4%), followed by model size (62.5%) and chunking strategy (60.8%), confirming that prompting, aggregation, and temperature exert comparatively minor influence.

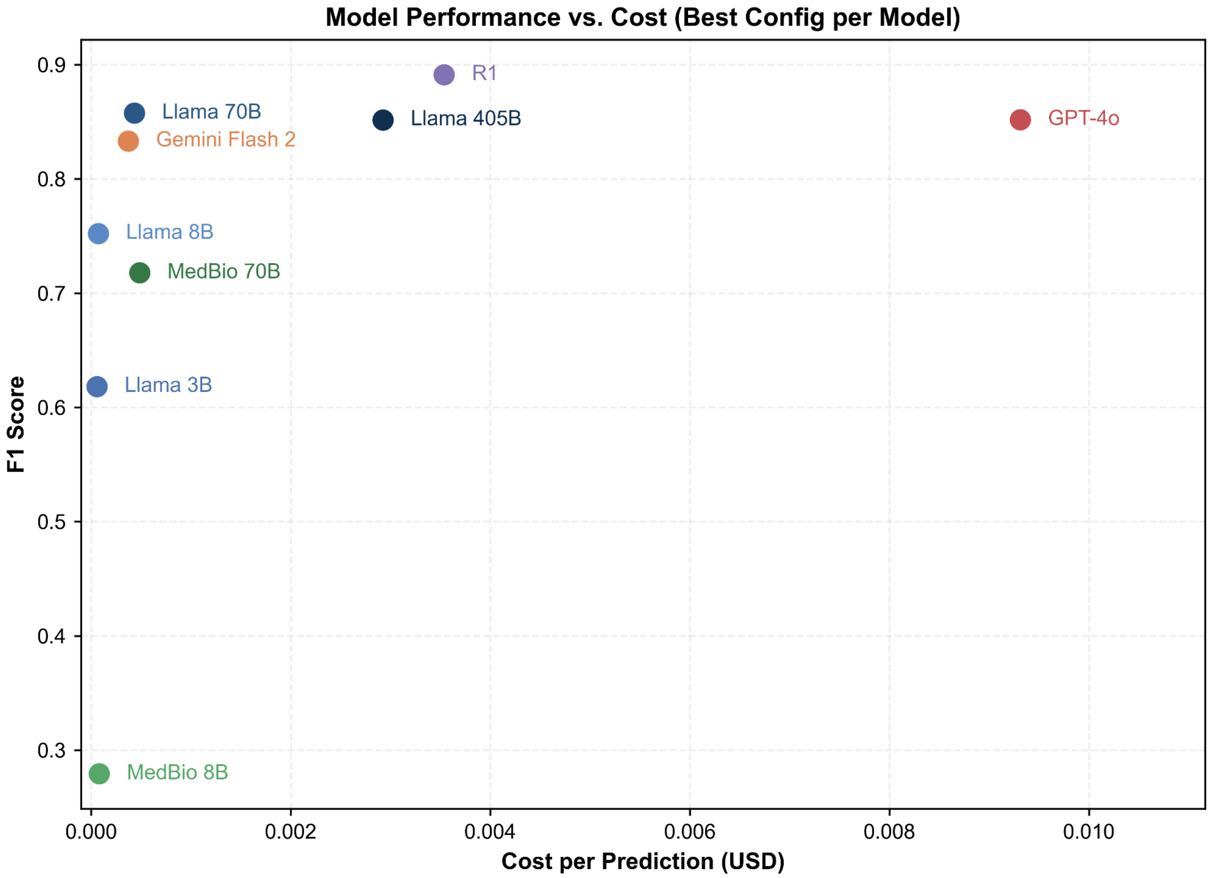


**Figure S6:** Cost–performance tradeoff across language models using best configuration per model

The plot shows a non-linear relationship between F1 score and cost, with early gains in performance occurring at minimal cost increases, followed by a plateau despite rising costs.

Each point represents a model's mean F1 score (y-axis) and cost per prediction (x-axis) using its best-performing configuration. Models are grouped by category: open-source models (e.g., LLaMA 3B–70B, MedBio 8B/70B) offer the lowest cost, with LLaMA 70B achieving performance (F1 = 0.85) on par with larger commercial and frontier models. Commercial efficient models (e.g., Gemini Flash 2.0) provide strong performance at moderate cost. Frontier models (e.g., LLaMA 405B, DeepSeek R1, GPT-4o) deliver the highest F1 scores ($\geq 0.84$) but incur the highest per-query costs.
