## Supplementary Tables for "A Systematic Exploration of LLM Behavior for EHR Phenotyping"

### 1 Supplementary Tables

**Table S1:** LLM performance for EHR phenotyping (best configuration for model category)

| Model Category | Model | Best Configuration | Precision | Recall | Accuracy | Macro F1 | Micro F1 | F1 efficiency |
|---|---|---|---|---|---|---|---|---|
| Baseline | LLaMa 3B | ZS+N | 0.697 (±0.273) | 0.535 (±0.229) | 0.866 (±0.102) | 0.534 (±0.166) | 0.556 | 0.638 |
| Open Source Models | LLaMa 3B | COT+N | 0.634 (±0.26) | 0.723 (±0.20) | 0.875 (±0.07) | 0.618 (±0.17) | 0.665 | 0.713 |
| | LLaMa 8B | ZS+N | 0.744 (±0.18) | 0.811 (±0.17) | 0.926 (±0.06) | 0.752 (±0.14) | 0.792 | 0.817 |
| | LLaMa 70B | ZS+N | 0.830 (±0.12) | 0.903 (±0.13) | 0.962 (±0.02) | 0.858 (±0.11) | 0.898 | **0.896** |
| | LLaMa 405B | COT+N | 0.819 (±0.12) | 0.906 (±0.16) | 0.960 (±0.03) | 0.852 (±0.13) | 0.892 | 0.841 |
| Fine-tuned Models | MedBio 8B | COT+N | 0.184 (±0.18) | **0.953 (±0.04)** | 0.251 (±0.15) | 0.279 (±0.21) | 0.313 | 0.429 |
| | MedBio 70B | COT+N | 0.626 (±0.26) | 0.916 (±0.13) | 0.905 (±0.06) | 0.718 (±0.18) | 0.777 | 0.780 |
| Commercial Models | GPT-4o | COT+N | 0.808 (±0.14) | 0.922 (±0.16) | 0.959 (±0.03) | 0.852 (±0.14) | 0.891 | 0.712 |
| | Gemini Flash 2.0.0 | COT+N | 0.760 (±0.09) | 0.937 (±0.14) | 0.948 (±0.03) | 0.833 (±0.11) | 0.87 | 0.877 |
| Reasoning Models | DeepSeekR1 | ZS+N | **0.883 (±0.09)** | 0.907 (±0.11) | **0.971 (±0.02)** | **0.891 (±0.09)** | **0.919** | 0.861 |

*Note.* ZS = Zero-Shot, CoT = Chain-of-Thought, N = No chunking.
Standard deviations (±) are shown next to means for all metrics except Micro F1 and F1 efficiency.
Bold values indicate the best performance for each metric.

**Table S2:** Complete Experimental Results for all models

| Model | Prompt | Chunking | Aggregation | Precision | Recall | Accuracy | Macro F1 | Micro F1 | F1 Efficiency |
|---|---|---|---|---|---|---|---|---|---|
| LLaMa 3B | Zero-Shot | None | N/A | 0.697 (±0.273) | 0.535 (±0.229) | 0.866 (±0.102) | 0.534 (±0.166) | 0.556 | 0.638 |
| LLaMa 3B | Zero-Shot | Document | Any | 0.477 (±0.237) | 0.855 (±0.106) | 0.824 (±0.086) | 0.571 (±0.209) | 0.634 | 0.668 |
| LLaMa 3B | Zero-Shot | Document | $\geq 2$ | 0.683 (±0.251) | 0.556 (±0.205) | 0.879 (±0.081) | 0.552 (±0.150) | 0.617 | 0.652 |
| LLaMa 3B | Zero-Shot | Document | Maj | 0.851 (±0.291) | 0.068 (±0.073) | 0.833 (±0.155) | 0.114 (±0.115) | 0.137 | 0.293 |
| LLaMa 3B | Zero-Shot | Naive | Any | 0.486 (±0.235) | 0.807 (±0.133) | 0.814 (±0.109) | 0.559 (±0.202) | 0.601 | 0.658 |
| LLaMa 3B | Zero-Shot | Naive | $\geq 2$ | 0.671 (±0.257) | 0.528 (±0.233) | 0.861 (±0.098) | 0.514 (±0.145) | 0.553 | 0.621 |
| LLaMa 3B | Zero-Shot | Naive | Maj | 0.829 (±0.174) | 0.138 (±0.172) | 0.835 (±0.165) | 0.199 (±0.209) | 0.169 | 0.363 |
| LLaMa 3B | Few-Shot | None | N/A | 0.464 (±0.254) | 0.890 (±0.106) | 0.788 (±0.154) | 0.565 (±0.240) | 0.596 | 0.663 |
| LLaMa 3B | Few-Shot | Document | Any | 0.262 (±0.211) | 0.983 (±0.020) | 0.498 (±0.221) | 0.377 (±0.230) | 0.412 | 0.509 |
| LLaMa 3B | Few-Shot | Document | $\geq 2$ | 0.375 (±0.251) | 0.885 (±0.116) | 0.686 (±0.212) | 0.472 (±0.222) | 0.498 | 0.587 |
| LLaMa 3B | Few-Shot | Document | Maj | 0.643 (±0.269) | 0.409 (±0.264) | 0.835 (±0.119) | 0.394 (±0.193) | 0.453 | 0.522 |
| LLaMa 3B | Few-Shot | Naive | Any | 0.287 (±0.221) | 0.973 (±0.035) | 0.552 (±0.215) | 0.403 (±0.234) | 0.435 | 0.53 |
| LLaMa 3B | Few-Shot | Naive | $\geq 2$ | 0.398 (±0.260) | 0.833 (±0.139) | 0.715 (±0.186) | 0.478 (±0.213) | 0.503 | 0.591 |
| LLaMa 3B | Few-Shot | Naive | Maj | 0.627 (±0.277) | 0.455 (±0.262) | 0.827 (±0.133) | 0.422 (±0.181) | 0.435 | 0.545 |
| LLaMa 3B | Chain-of-Thought | None | N/A | 0.634 (±0.260) | 0.723 (±0.196) | 0.875 (±0.073) | 0.618 (±0.173) | 0.665 | 0.707 |
| LLaMa 3B | Chain-of-Thought | Document | Any | 0.489 (±0.262) | 0.807 (±0.159) | 0.798 (±0.158) | 0.548 (±0.209) | 0.589 | 0.649 |
| LLaMa 3B | Chain-of-Thought | Document | $\geq 2$ | 0.666 (±0.269) | 0.526 (±0.239) | 0.850 (±0.111) | 0.499 (±0.163) | 0.544 | 0.608 |
| LLaMa 3B | Chain-of-Thought | Document | Maj | 0.623 (±0.417) | 0.071 (±0.139) | 0.831 (±0.164) | 0.104 (±0.166) | 0.124 | 0.284 |
| LLaMa 3B | Chain-of-Thought | Naive | Any | 0.500 (±0.268) | 0.792 (±0.168) | 0.801 (±0.137) | 0.548 (±0.209) | 0.581 | 0.649 |
| LLaMa 3B | Chain-of-Thought | Naive | $\geq 2$ | 0.655 (±0.267) | 0.520 (±0.251) | 0.852 (±0.102) | 0.486 (±0.151) | 0.53 | 0.598 |
| LLaMa 3B | Chain-of-Thought | Naive | Maj | 0.769 (±0.256) | 0.127 (±0.188) | 0.834 (±0.168) | 0.184 (±0.197) | 0.167 | 0.35 |
| LLaMa 3B | ETP | None | ETP | 0.494 (±0.251) | 0.792 (±0.189) | 0.822 (±0.092) | 0.547 (±0.184) | 0.618 | 0.775 |

| Model | Prompt | Chunking | Aggregation | Precision | Recall | Accuracy | Macro F1 | Micro F1 | F1 Efficiency |
|---|---|---|---|---|---|---|---|---|---|
| LLaMa 3B | ETP | Document | ETP | 0.217 (±0.199) | 0.833 (±0.343) | 0.542 (±0.266) | 0.318 (±0.235) | 0.398 | 0.458 |
| LLaMa 3B | ETP | Naive | ETP | 0.230 (±0.207) | 0.826 (±0.342) | 0.580 (±0.244) | 0.331 (±0.240) | 0.415 | 0.469 |
| LLaMa 8B | Zero-Shot | None | N/A | 0.744 (±0.181) | 0.811 (±0.170) | 0.926 (±0.063) | 0.752 (±0.144) | 0.792 | 0.817 |
| LLaMa 8B | Zero-Shot | Document | Any | 0.478 (±0.187) | 0.960 (±0.046) | 0.844 (±0.067) | 0.614 (±0.176) | 0.687 | 0.702 |
| LLaMa 8B | Zero-Shot | Document | $\geq 2$ | 0.699 (±0.203) | 0.748 (±0.143) | 0.910 (±0.051) | 0.692 (±0.128) | 0.751 | 0.766 |
| LLaMa 8B | Zero-Shot | Document | Maj | 0.932 (±0.101) | 0.152 (±0.101) | 0.852 (±0.132) | 0.248 (±0.146) | 0.302 | 0.402 |
| LLaMa 8B | Zero-Shot | Naive | Any | 0.493 (±0.195) | 0.949 (±0.066) | 0.849 (±0.067) | 0.623 (±0.181) | 0.691 | 0.709 |
| LLaMa 8B | Zero-Shot | Naive | $\geq 2$ | 0.690 (±0.202) | 0.750 (±0.159) | 0.904 (±0.061) | 0.683 (±0.126) | 0.732 | 0.759 |
| LLaMa 8B | Zero-Shot | Naive | Maj | 0.887 (±0.138) | 0.279 (±0.189) | 0.861 (±0.138) | 0.391 (±0.209) | 0.384 | 0.52 |
| LLaMa 8B | Few-Shot | None | N/A | 0.547 (±0.229) | 0.916 (±0.091) | 0.870 (±0.063) | 0.651 (±0.199) | 0.712 | 0.734 |
| LLaMa 8B | Few-Shot | Document | Any | 0.285 (±0.190) | 0.990 (±0.008) | 0.598 (±0.146) | 0.413 (±0.208) | 0.468 | 0.537 |
| LLaMa 8B | Few-Shot | Document | $\geq 2$ | 0.415 (±0.214) | 0.895 (±0.053) | 0.786 (±0.092) | 0.534 (±0.209) | 0.6 | 0.636 |
| LLaMa 8B | Few-Shot | Document | Maj | 0.762 (±0.213) | 0.373 (±0.147) | 0.873 (±0.105) | 0.470 (±0.144) | 0.514 | 0.584 |
| LLaMa 8B | Few-Shot | Naive | Any | 0.303 (±0.195) | 0.987 (±0.017) | 0.627 (±0.157) | 0.433 (±0.209) | 0.485 | 0.554 |
| LLaMa 8B | Few-Shot | Naive | $\geq 2$ | 0.431 (±0.224) | 0.880 (±0.087) | 0.787 (±0.101) | 0.538 (±0.204) | 0.591 | 0.64 |
| LLaMa 8B | Few-Shot | Naive | Maj | 0.714 (±0.239) | 0.446 (±0.184) | 0.869 (±0.112) | 0.505 (±0.148) | 0.527 | 0.613 |
| LLaMa 8B | Chain-of-Thought | None | N/A | 0.677 (±0.208) | 0.841 (±0.158) | 0.909 (±0.065) | 0.720 (±0.153) | 0.757 | 0.79 |
| LLaMa 8B | Chain-of-Thought | Document | Any | 0.528 (±0.215) | 0.916 (±0.101) | 0.861 (±0.065) | 0.634 (±0.187) | 0.693 | 0.718 |
| LLaMa 8B | Chain-of-Thought | Document | $\geq 2$ | 0.741 (±0.209) | 0.667 (±0.174) | 0.898 (±0.091) | 0.660 (±0.147) | 0.679 | 0.739 |
| LLaMa 8B | Chain-of-Thought | Document | Maj | 0.935 (±0.117) | 0.092 (±0.063) | 0.835 (±0.163) | 0.160 (±0.098) | 0.147 | 0.329 |
| LLaMa 8B | Chain-of-Thought | Naive | Any | 0.561 (±0.225) | 0.893 (±0.119) | 0.865 (±0.075) | 0.647 (±0.189) | 0.691 | 0.729 |
| LLaMa 8B | Chain-of-Thought | Naive | $\geq 2$ | 0.738 (±0.221) | 0.656 (±0.196) | 0.890 (±0.098) | 0.644 (±0.149) | 0.649 | 0.727 |
| LLaMa 8B | Chain-of-Thought | Naive | Maj | 0.898 (±0.116) | 0.196 (±0.179) | 0.843 (±0.164) | 0.288 (±0.197) | 0.24 | 0.434 |

| Model | Prompt | Chunking | Aggregation | Precision | Recall | Accuracy | Macro F1 | Micro F1 | F1 Efficiency |
|---|---|---|---|---|---|---|---|---|---|
| LLaMa 8B | ETP | None | ETP | 0.688 (±0.174) | 0.804 (±0.218) | 0.909 (±0.057) | 0.701 (±0.141) | 0.772 | 0.775 |
| LLaMa 8B | ETP | Document | ETP | 0.384 (±0.226) | 0.790 (±0.338) | 0.817 (±0.108) | 0.496 (±0.241) | 0.614 | 0.605 |
| LLaMa 8B | ETP | Naive | ETP | 0.375 (±0.218) | 0.790 (±0.338) | 0.816 (±0.098) | 0.489 (±0.237) | 0.614 | 0.599 |
| LLaMa 70B | Zero-Shot | None | N/A | 0.830 (±0.122) | 0.903 (±0.131) | 0.962 (±0.023) | 0.858 (±0.108) | 0.898 | 0.896 |
| LLaMa 70B | Zero-Shot | Document | Any | 0.517 (±0.242) | 0.958 (±0.069) | 0.852 (±0.102) | 0.638 (±0.233) | 0.701 | 0.711 |
| LLaMa 70B | Zero-Shot | Document | $\geq 2$ | 0.683 (±0.217) | 0.794 (±0.143) | 0.911 (±0.052) | 0.705 (±0.151) | 0.767 | 0.765 |
| LLaMa 70B | Zero-Shot | Document | Maj | 0.935 (±0.111) | 0.253 (±0.129) | 0.868 (±0.119) | 0.378 (±0.170) | 0.424 | 0.497 |
| LLaMa 70B | Zero-Shot | Naive | Any | 0.559 (±0.230) | 0.963 (±0.053) | 0.876 (±0.098) | 0.677 (±0.215) | 0.737 | 0.743 |
| LLaMa 70B | Zero-Shot | Naive | $\geq 2$ | 0.724 (±0.207) | 0.791 (±0.141) | 0.921 (±0.048) | 0.729 (±0.143) | 0.784 | 0.785 |
| LLaMa 70B | Zero-Shot | Naive | Maj | 0.923 (±0.101) | 0.333 (±0.177) | 0.875 (±0.121) | 0.464 (±0.191) | 0.477 | 0.568 |
| LLaMa 70B | Few-Shot | None | N/A | 0.794 (±0.163) | 0.925 (±0.108) | 0.954 (±0.040) | 0.843 (±0.129) | 0.88 | 0.883 |
| LLaMa 70B | Few-Shot | Document | Any | 0.508 (±0.223) | 0.973 (±0.040) | 0.843 (±0.121) | 0.637 (±0.222) | 0.691 | 0.707 |
| LLaMa 70B | Few-Shot | Document | $\geq 2$ | 0.671 (±0.209) | 0.820 (±0.127) | 0.906 (±0.064) | 0.710 (±0.157) | 0.763 | 0.767 |
| LLaMa 70B | Few-Shot | Document | Maj | 0.918 (±0.107) | 0.268 (±0.136) | 0.871 (±0.115) | 0.394 (±0.165) | 0.448 | 0.508 |
| LLaMa 70B | Few-Shot | Naive | Any | 0.554 (±0.217) | 0.975 (±0.051) | 0.870 (±0.110) | 0.680 (±0.208) | 0.731 | 0.742 |
| LLaMa 70B | Few-Shot | Naive | $\geq 2$ | 0.710 (±0.206) | 0.806 (±0.125) | 0.917 (±0.055) | 0.730 (±0.151) | 0.78 | 0.784 |
| LLaMa 70B | Few-Shot | Naive | Maj | 0.905 (±0.117) | 0.349 (±0.175) | 0.878 (±0.117) | 0.479 (±0.182) | 0.498 | 0.577 |
| LLaMa 70B | Chain-of-Thought | None | N/A | 0.825 (±0.123) | 0.901 (±0.153) | 0.962 (±0.023) | 0.854 (±0.124) | 0.896 | 0.893 |
| LLaMa 70B | Chain-of-Thought | Document | Any | 0.525 (±0.212) | 0.968 (±0.035) | 0.862 (±0.092) | 0.654 (±0.195) | 0.717 | 0.722 |
| LLaMa 70B | Chain-of-Thought | Document | $\geq 2$ | 0.701 (±0.188) | 0.800 (±0.132) | 0.915 (±0.051) | 0.724 (±0.127) | 0.778 | 0.779 |
| LLaMa 70B | Chain-of-Thought | Document | Maj | 0.939 (±0.079) | 0.258 (±0.124) | 0.868 (±0.121) | 0.388 (±0.161) | 0.428 | 0.504 |
| LLaMa 70B | Chain-of-Thought | Naive | Any | 0.577 (±0.207) | 0.968 (±0.052) | 0.889 (±0.084) | 0.699 (±0.187) | 0.759 | 0.759 |
| LLaMa 70B | Chain-of-Thought | Naive | $\geq 2$ | 0.740 (±0.170) | 0.787 (±0.151) | 0.925 (±0.045) | 0.743 (±0.121) | 0.792 | 0.795 |

| Model | Prompt | Chunking | Aggregation | Precision | Recall | Accuracy | Macro F1 | Micro F1 | F1 Efficiency |
|---|---|---|---|---|---|---|---|---|---|
| LLaMa 70B | Chain-of-Thought | Naive | Maj | 0.926 (±0.101) | 0.333 (±0.187) | 0.874 (±0.125) | 0.462 (±0.203) | 0.471 | 0.565 |
| LLaMa 70B | ETP | None | ETP | 0.763 (±0.169) | 0.840 (±0.144) | 0.931 (±0.048) | 0.783 (±0.126) | 0.819 | 0.834 |
| LLaMa 70B | ETP | Document | ETP | 0.552 (±0.274) | 0.773 (±0.326) | 0.897 (±0.071) | 0.631 (±0.281) | 0.733 | 0.699 |
| LLaMa 70B | ETP | Naive | ETP | 0.554 (±0.278) | 0.783 (±0.330) | 0.898 (±0.073) | 0.635 (±0.285) | 0.736 | 0.702 |
| LLaMa 405B | Chain-of-Thought | None | N/A | 0.819 (±0.124) | 0.906 (±0.161) | 0.960 (±0.027) | 0.852 (±0.133) | 0.892 | 0.841 |
| MedBio 8B | Chain-of-Thought | None | N/A | 0.184 (±0.177) | 0.953 (±0.035) | 0.251 (±0.153) | 0.279 (±0.205) | 0.313 | 0.429 |
| MedBio 70B | Chain-of-Thought | None | N/A | 0.626 (±0.215) | 0.916 (±0.128) | 0.905 (±0.059) | 0.718 (±0.182) | 0.777 | 0.78 |
| GPT-4o | Chain-of-Thought | None | N/A | 0.808 (±0.136) | 0.922 (±0.164) | 0.959 (±0.031) | 0.852 (±0.144) | 0.891 | 0.713 |
| Gemini Flash | Chain-of-Thought | None | N/A | 0.760 (±0.095) | 0.937 (±0.142) | 0.948 (±0.033) | 0.833 (±0.107) | 0.87 | 0.877 |
| R1 | Zero-Shot | None | N/A | 0.883 (±0.094) | 0.907 (±0.108) | 0.971 (±0.023) | 0.891 (±0.091) | 0.919 | 0.861 |
