## Supplementary material for "A Systematic Exploration of LLM Behavior for EHR Phenotyping": S2 LLM Prompts

### 1 Supplemental File 2 - LLM Prompts

#### System prompt

```
SYSTEM_PROMPT = """
You are a medical researcher specializing in analyzing discharge summaries for phenotype
classification in epidemiologic studies.
Your task is to determine the presence or absence of a given condition based on the available
evidence in the discharge summary.

Guidelines:
- Thoroughly examine the discharge summary for all relevant medical information.
- Prioritize documented clinical evidence over speculation.
- Interpret standard medical abbreviations and terminologies accurately.

Classification approach:
- Make decisive classifications based on the strongest available evidence.
- For uncertain cases, favor classifications supported by evidence.
- Maintain high accuracy, as your classifications will directly impact the study's validity.
"""
```

#### User prompts

```
PROMPT_ZERO_SHOT = """
Analyze the discharge summary or a specific segment of it provided below to determine the presence
of {phenotype}.
Use your professional judgment to critically assess the information presented and determine if the
history, symptoms and treatments described indicate that the patient has {phenotype}.
It is critical to ensure accuracy in your analysis, as errors can have significant consequences.
Conclude with a JSON object stating the status of {phenotype} presence.
Only output 1 if the evidence is conclusive.

Discharge Summary or Segment to Analyze:
{discharge_summary}

Assessment Instructions:
1. Review all clinical information.
2. Determine whether there is conclusive evidence supporting a diagnosis of **{phenotype}**.
   - If conclusive evidence exists, respond with "status": "1"
   - If no conclusive evidence exists, respond with "status": "0"

- Your response must only contain a JSON object on a new line.
- Do not provide any explanations or additional text.
- The JSON object should follow this exact format:
{{"phenotype": "{phenotype}", "status": "1 or 0"}}
"""




PROMPT_FEW_SHOT = """
Analyze the discharge summary or a specific segment of it provided below to determine the presence
of {phenotype}.
```

```
Use your professional judgment to critically assess the information presented and determine if the
history, symptoms and treatments described indicate that the patient has {phenotype}.
It is critical to ensure accuracy in your analysis, as errors can have significant consequences.
Only output 1 if the evidence is conclusive.

Examples:
- Discharge Summary: "Patient was admitted following a heart attack and underwent angioplasty."
  {{"phenotype": "Myocardial Infarction", "status": "1"}}
- Discharge Summary: "The patient presented with new onset hemiplegia, brain MRI was found normal
but cervical MRI showed transverse myelitis"
  {{"phenotype": "Stroke", "status": "0"}}
- Discharge Summary: "The patient past medical history is relevant for VTE."
  {{"phenotype": "Deep Venous thrombosis", "status": "1"}}

Discharge Summary or Segment to Analyze:
{discharge_summary}

Instructions:
1. Review all clinical information in the provided discharge summary.
2. Determine if there is conclusive evidence supporting a diagnosis of {phenotype}.
3. Respond with a JSON object containing only the phenotype and status.
4. Use "status": "1" if {phenotype} is conclusively present, "status": "0" if not.

- Your response must contain ONLY the JSON object on a new line.
- Do not provide any explanations or additional text.

OUTPUT:
{{"phenotype": "{phenotype}", "status": "1 or 0"}}
"""


PROMPT_COT = """
Analyze the discharge summary provided below to determine the presence of {phenotype}.
Use a detailed reasoning process as a medical doctor would to arrive at your conclusion.
It is imperative that your response be accurate, as errors cannot be tolerated.

Discharge Summary:
{discharge_summary}

Step-by-Step Reasoning:
1. Identify and list the key features mentioned in the discharge summary that are relevant to {
phenotype}.
2. Assess whether these features align more with the presence or absence of {phenotype}. Only
conclude presence if the evidence is conclusive.
3. Summarize your findings based on the evidence and make a professional judgment.

Instructions:
- Provide a JSON object with your final judgment, including a classification status and a
justification.
- Use "status": "1" if {phenotype} is conclusively present, "status": "0" if not.
- Limit your justification to approximately 50 tokens.
- Your response must contain ONLY the JSON object, starting on a new line.
- Do not provide any explanations or additional text outside the JSON object.

{{"phenotype": "{phenotype}", "status": "1 or 0", "justification": "Your 50-token Justification
Here"}}
"""
```

```
PROMPT_COT_CONFIDENCE = """
Analyze the discharge summary provided below to determine the presence of {phenotype}.
Use a detailed reasoning process as a medical doctor would to arrive at your conclusion.
It is imperative that your response be accurate, as errors cannot be tolerated.

Discharge Summary:
{discharge_summary}

Step-by-Step Reasoning:
1. Identify and list the key features mentioned in the discharge summary that are relevant to {
phenotype}.
2. Assess whether these features align more with the presence or absence of {phenotype}. Only
conclude presence if the evidence is conclusive.
3. Summarize your findings based on the evidence and make a professional judgment.
4. Rate your personal confidence in this conclusion (high/medium/low).
- High confidence: Clear diagnostic criteria met
- Medium confidence: Suggestive features with minimal ambiguity
- Low confidence: Equivocal/incomplete information

Instructions:
- Provide a JSON object with your final judgment, including a classification status, justification
and confidence level.
- Use "status": "1" if {phenotype} is conclusively present, "status": "0" if not.
- Limit your justification to approximately 50 tokens.
- Include a confidence level of "high", "medium", or "low" based on your certainty in the
prediction.
- Your response must contain ONLY the JSON object, starting on a new line.
- Do not provide any explanations or additional text outside the JSON object.

{{"phenotype": "{phenotype}", "status": "1 or 0", "justification": "Your 50-token Justification
Here", "confidence": "high/medium/low"}}
"""
```

#### EXTRACT-THEN-PHENOTYTPE-PROMPT

```
PROMPT_ETP_DMII = '''
Analyze the provided discharge summary segment to determine the presence of {phenotype} Type II
diabetes and associated details.
Use your judgment to assess symptoms, treatments, and complications indicative of {phenotype} Type
II diabetes.

Discharge Summary or Segment to Analyze:
{discharge_summary}

Instructions:
Determine if any of the following elements related to {phenotype} are present in the segment:
- '{phenotype}': Explicit positive mention of Type II diabetes.
- 'metformin', 'sulfonylureas', 'glp-1 agonists', 'dpp-4 inhibitors', 'sglt2 inhibitors', 'insulin
': Medication status.
  - Non exhaustive list of medications corresponding to those categories:
    - Metformin: Metformin, Glucophage
    - Sulfonylureas: Glipizide, Glyburide, Glimepiride
    - GLP-1 Agonists: Exenatide (Byetta), Liraglutide (Victoza), Dulaglutide (Trulicity)
    - DPP-4 Inhibitors: Sitagliptin (Januvia), Saxagliptin (Onglyza), Linagliptin (Tradjenta)
    - SGLT2 Inhibitors: Canagliflozin (Invokana), Dapagliflozin (Farxiga), Empagliflozin (Jardiance
)
```

```
    - Insulin: Humulin, Novolin, Lantus, Levemir, etc.
- 'neuropathy', 'retinopathy', 'nephropathy': Presence of diabetes-related complications
- 'hba1c': Mention of HbA1c levels >= 6.5%, a diagnostic criterion for Type II diabetes.
- 'glucose_levels': Venous or capillary glucose > 200 mg/dL


For ambiguous cases or partial mentions, use your best judgment to determine if the element is
sufficiently indicative of {phenotype} Type II diabetes.

Conclude with a JSON object that states the status of each predefined element. Respond with '1' for
 present and '0' for absent.
The keys should include:

OUTPUT:
{{
  "{phenotype}": "1 or 0",
  "metformin": "1 or 0",
  "sulfonylureas": "1 or 0",
  "glp1_agonists": "1 or 0",
  "dpp4_inhibitors": "1 or 0",
  "sglt2_inhibitors": "1 or 0",
  "insulin": "1 or 0",
  "neuropathy": "1 or 0",
  "retinopathy": "1 or 0",
  "nephropathy": "1 or 0",
  "hba1c": "1 or 0",
  "gluose_levels": "1 or 0"
}}
'''

PROMPT_ETP_DMI = '''
Analyze the provided discharge summary segment to determine the presence of {phenotype} Type I
diabetes and associated details.
Use your judgment to assess symptoms, treatments, and complications indicative of {phenotype}.

Discharge Summary or Segment to Analyze:
{discharge_summary}

Instructions:
Determine if any of the following elements related to {phenotype} are present in the segment:
- '{phenotype}': Explicit positive mention of Type I diabetes.
- 'insulin_therapy_from_diagnosis': Dependence on insulin from the time of diagnosis.
- 'insulin_therapy_any': General mention of insulin therapy.
- 'autoantibodies': Presence of specific anti-islets antibodies such as anti-GAD, anti-ZnT8, anti-
IA2, and anti-Insulin antibodies, which support a diagnosis of {phenotype}.
- 'ketoacidosis': Previous or current diabetic ketoacidosis. Synonyms: DKA, diabetic ketoacidosis.
- 'no_oral_hypoglycemics': Absence of oral hypoglycemic agents.
- 'neuropathy', 'retinopathy', 'nephropathy': Complications presence.

Conclude with a JSON object that states the status of each predefined element. Respond with '1' for
 present and '0' for absent.
The keys should include:

OUTPUT:
{{
  "{phenotype}": "1 or 0",
  "insulin_therapy_from_diagnosis": "1 or 0",
  "insulin_therapy_any": "1 or 0",
  "autoantibodies": "1 or 0",
```

```
  "ketoacidosis": "1 or 0",
  "no_oral_hypoglycemics": "1 or 0",
  "neuropathy": "1 or 0",
  "retinopathy": "1 or 0",
  "nephropathy": "1 or 0",
}}
'''

PROMPT_ETP_CDIFF_PAST = '''
Analyze the provided discharge summary to determine if there is any past medical history of C.
difficile infection prior to the current admission.
Use your judgment to assess historical documentation of prior CDI episodes.

Discharge Summary to Analyze:
{discharge_summary}

Instructions:
Identify these historical CDI-related elements from before the current admission:
- '{phenotype}': Explicit mention of previous episodes of C. difficile, C. diff, CDI, or
pseudomembranous colitis
- 'recurrent': Documentation of recurrent or multiple past episodes
- 'prior_treatment': Previous treatment with oral vancomycin, fidaxomicin, or metronidazole
- 'prior_fmt': Previous fecal microbiota transplantation
- 'prior_complications': Past complications from CDI (e.g., toxic megacolon, colectomy)

Conclude with a JSON object that states the status of each predefined element. Respond with '1' for
 present and '0' for absent.

OUTPUT:
{{
  "{phenotype}": "1 or 0",
  "recurrent": "1 or 0",
  "prior_treatment": "1 or 0",
  "prior_fmt": "1 or 0",
  "prior_complications": "1 or 0"
}}
'''

PROMPT_ETP_CDIFF_COMPLICATION = '''
Analyze the provided discharge summary to determine if C. difficile infection occurred as a
complication during the current admission.
Use your judgment to assess new onset CDI during this hospitalization.

Discharge Summary to Analyze:
{discharge_summary}

Instructions:
Identify these CDI-related elements that developed during the current admission:
- '{phenotype}': Explicit mention of new diagnosis of C. difficile, C. diff, CDI, or
pseudomembranous colitis
- 'symptoms': New onset severe diarrhea, abdominal pain/cramping, fever
- 'diagnostic_tests': Positive C. difficile toxin test, PCR, or GDH assay during this admission
- 'colonoscopy': Pseudomembranes or other CDI findings on colonoscopy this admission
- 'treatment': Active use of oral vancomycin, fidaxomicin, or metronidazole for CDI treatment
- 'complications': Development of complications (toxic megacolon, shock, colectomy)
- 'fmt_needed': Requirement for fecal microbiota transplantation
- 'nosocomial': Clear evidence infection developed >48 hours after admission
```

```
Conclude with a JSON object that states the status of each predefined element. Respond with '1' for
 present and '0' for absent.

OUTPUT:
{{
  "{phenotype}": "1 or 0",
  "symptoms": "1 or 0",
  "diagnostic_tests": "1 or 0",
  "colonoscopy": "1 or 0",
  "treatment": "1 or 0",
  "complications": "1 or 0",
  "fmt_needed": "1 or 0",
  "nosocomial": "1 or 0"
}}
'''

PROMPT_ETP_DEMENTIA = '''
Analyze the provided discharge summary segment to determine the presence of {phenotype} and
associated details.
Use your judgment to assess symptoms, treatments, and diagnostic indicators indicative of {
phenotype}.

Discharge Summary or Segment to Analyze:
{discharge_summary}

Instructions:
Determine if any of the following elements related to {phenotype} are present in the segment:
- '{phenotype}': Explicit mention of neurodegenerative diseases or related terms, including but not
 limited to:
  - Dementia (including variants like vascular dementia, Parkinson's, Lewy body dementia, Alzheimer
's disease, Frontotemporal dementia, etc.)
    But not Mild cognitive impairment (MCI).
- 'cognitive_decline': Evidence of progressive cognitive decline
- 'memory_loss': Frequent or severe memory loss, especially short-term memory loss.
- 'disorientation': Disorientation to time and place.
- 'behavioral_changes': Changes in behavior or personality.
- 'cholinesterase_inhibitors': Use of medications like donepezil, rivastigmine. Synonyms: Aricept,
Exelon.
- 'memantine': Use of memantine. Synonyms: Namenda.
- 'neuroimaging_findings': MRI or CT findings that support a diagnosis of {phenotype}, such as
atrophy in specific brain areas.
- 'functional_decline': Loss of ability to perform daily activities independently. Synonyms: loss
of independence, functional impairment.

Conclude with a JSON object that states the status of each predefined element. Respond with '1' for
 present and '0' for absent.

The keys should include:

OUTPUT:
{{
  "{phenotype}": "1 or 0",
  "cognitive_decline": "1 or 0",
  "memory_loss": "1 or 0",
  "disorientation": "1 or 0",
  "behavioral_changes": "1 or 0",
  "cholinesterase_inhibitors": "1 or 0",
  "memantine": "1 or 0",
  "neuroimaging_findings": "1 or 0",
```

```
  "functional_decline": "1 or 0"
}}
'''

PROMPT_ETP_DEPRESSION = '''
Analyze the provided discharge summary segment to determine the presence of {phenotype} and
associated details.
Use your clinical judgment to assess symptoms, treatments, and diagnostic indicators indicative of
{phenotype}.

Discharge Summary or Segment to Analyze:
{discharge_summary}

Instructions:
Determine if any of the following elements related to {phenotype} are present in the segment:
- '{phenotype}': Explicit mention of major depressive disorder or bipolar disorder.
- 'fatigue': Reports of significant fatigue or loss of energy nearly every day.
- 'mood_symptoms': Sadness, emptiness, anhedonia, loss of interest/pleasure. Synonyms: chronic
sadness, feelings of emptiness, lack of enjoyment, loss of interest.
- 'sleep_disturbances': Changes in sleeping patterns, either insomnia or hypersomnia, associated
with {phenotype}.
- 'appetite_changes': Significant weight loss or gain due to changes in appetite, common in {
phenotype}.
- 'antidepressants': Use of specific medications, including but not limited to:
  - SSRIs (e.g., fluoxetine (Prozac), sertraline (Zoloft), paroxetine (Paxil))
  - SNRIs (e.g., venlafaxine (Effexor), duloxetine (Cymbalta))
  - TCAs (e.g., amitriptyline (Elavil), imipramine (Tofranil))
  - MAOIs (e.g., phenelzine (Nardil), tranylcypromine (Parnate))
  - Other antidepressants (e.g., bupropion (Wellbutrin), mirtazapine (Remeron))
- 'psychotherapy': Engagement in psychotherapy sessions as part of treatment.
- 'suicide_risk': Suicidal thoughts, plans, or attempts. Synonyms: suicidal ideation, suicide
attempts.

Conclude with a JSON object that states the status of each predefined element. Respond with '1' for
 present and '0' for absent.
The keys should include:

OUTPUT:
{{
  "{phenotype}": "1 or 0",
  "mood_symptoms": "1 or 0",
  "sleep_disturbances": "1 or 0",
  "appetite_changes": "1 or 0",
  "antidepressants": "1 or 0",
  "psychotherapy": "1 or 0",
  "fatigue": "1 or 0",
  "suicide_risk": "1 or 0"
}}
'''

PROMPT_ETP_HTN = '''
Analyze the provided discharge summary segment to determine the presence of {phenotype} (
hypertension) and associated details.
Use your clinical judgment to assess risk factors, treatments, complications, and other indicators
of {phenotype}.

Discharge Summary or Segment to Analyze:
{discharge_summary}
```

```
Instructions:
Determine if any of the following elements related to {phenotype} are present in the segment:
- '{phenotype}': Explicit mention of hypertension or high blood pressure.
- 'blood_pressure_readings': Documented high blood pressure readings (e.g., systolic 130 mmHg or
diastolic 80 mmHg), or description of blood pressure as "elevated" or "high".
- 'antihypertensive_medications': Use of specific medications, including but not limited to:
  - ACE inhibitors (e.g., lisinopril, enalapril)
  - ARBs (e.g., losartan, valsartan)
  - Beta-blockers (e.g., metoprolol, atenolol)
  - Calcium channel blockers (e.g., amlodipine, diltiazem)
  - Thiazide diuretics (e.g., hydrochlorothiazide)
  - Other antihypertensives (e.g., clonidine, hydralazine)
- 'end-organ_damage': Evidence of hypertension-related complications affecting major organs:
     - Renal: Impaired kidney function
     - Ocular: Hypertensive retinopathy
     - Cardiac: Left ventricular hypertrophy, heart failure
     - Vascular: Coronary artery disease, peripheral artery disease
     - Cerebral: Stroke, transient ischemic attack (TIA)

Conclude with a JSON object that states the status of each predefined element. Respond with '1' for
 present and '0' for absent.
The keys should include:

OUTPUT:
{{
  "{phenotype}": "1 or 0",
  "blood pressure_readings": "1 or 0",
  "antihypertensive_medications": "1 or 0",
  "end-organ_damage": "1 or 0"
}}
'''

PROMPT_ETP_HFpEF = '''
Analyze the provided discharge summary segment to determine the presence of Heart Failure with
Preserved Ejection Fraction (HFpEF) and associated details.
Use your judgment to assess clinical signs, treatments, and diagnostic criteria indicative of {
phenotype}, ensuring distinction from HFrEF.

Discharge Summary or Segment to Analyze:
{discharge_summary}

Instructions:
Determine if any of the following elements related to {phenotype} are present in the segment:
- '{phenotype}': Explicit mention of Heart Failure with Preserved Ejection Fraction. Synonyms:
HFpEF, diastolic heart failure.
- 'preserved_EF': EF >50% or described as preserved/normal.
- 'diastolic_dysfunction': Mentioned in echocardiography or other cardiac imaging.
- 'congestion_symptoms': Dyspnea, orthopnea, edema.
- 'elevated_filling_pressures': E/e' ratio >14, elevated LAP or LVEDP mention on echocardiography
report.
- 'bnp_levels': Elevated NT-proBNP levels (> 900 pg/mL) or BNP levels (> 100 pg/mL)
- 'diuretics': Use of diuretics to manage fluid retention specifically furosemide (Lasix) in the
context of {phenotype}.

Conclude with a JSON object that states the status of each predefined element. Respond with '1' for
 present and '0' for absent.
The keys should include:

OUTPUT:
```

```
{{
  "{phenotype}": "1 or 0",
  "preserved_EF": "1 or 0",
  "diastolic_dysfunction": "1 or 0",
  "congestion_symptoms": "1 or 0",
  "elevated_filling_pressures": "1 or 0",
  "bnp_levels": "1 or 0",
  "diuretics": "1 or 0"
}}
'''


PROMPT_ETP_HFrEF = '''
Analyze the provided discharge summary segment to determine the presence of Heart Failure with
Reduced Ejection Fraction (HFrEF) and associated details.
Use your judgment to assess clinical signs, treatments, and diagnostic criteria indicative of {
phenotype}, ensuring clear differentiation from other forms of heart failure.

Discharge Summary or Segment to Analyze:
{discharge_summary}

Instructions:
Determine if any of the following elements related to {phenotype} are present in the segment:
- '{phenotype}': Explicit mention of Heart Failure with Reduced Ejection Fraction.
- 'reduced_EF': EF <= 40% or described as reduced/low systolic function.
- 'congestion_symptoms': Dyspnea, orthopnea, edema.
- 'bnp_levels': Elevated NT-proBNP levels (> 900 pg/mL) or BNP levels (> 100 pg/mL)
- 'beta-blockers': Use of beta-blockers (e.g., metoprolol, carvedilol).
- 'arni': Use of angiotensin receptor-neprilysin inhibitors (Entresto).
- 'diuretics': Use of loop diuretics (furosemide, torsemide, bumetanide, etc.)
- 'acei_arb': Prescription of ACE inhibitors or ARBs (example drugs: lisinopril, enalapril,
losartan).
- 'aldosterone_antagonists': Prescription of aldosterone antagonists (example drugs: spironolactone
 (aldactone), eplerenone).

Conclude with a JSON object that states the status of each predefined element. Respond with '1' for
 present and '0' for absent.
The keys should include:

OUTPUT:
{{
  "{phenotype}": "1 or 0",
  "reduced_EF": "1 or 0",
  "congestion_symptoms": "1 or 0",
  "bnp_levels": "1 or 0",
  "beta_blockers": "1 or 0",
  "arni": "1 or 0",
  "diuretics": "1 or 0",
  "acei_arb": "1 or 0",
  "aldosterone_antagonists": "1 or 0"
}}
'''


PROMPT_ETP_LUPUS = '''
Analyze the discharge summary segment for evidence of systemic lupus erythematosus (SLE) and not
simply cutaneous lupus.

Discharge Summary Segment:
```

```
{discharge_summary}

Instructions:
Identify the presence of these SLE-related elements:
- '{phenotype}': Explicit mention of SLE or systemic lupus erythematosus
- 'ANA': Positive antinuclear antibodies test (titer < 1:180)
- 'specific_antibodies': Positive Anti-dsDNA, anti-Sm, anti-phospholipid antibodies
- 'complement_levels': Low C3 or C4 complement levels
- 'skin_manifestations': Specific cutaneous manifestations of systemic lupus: malar rash, discoid
rash, photosensitivity
- 'joint_symptoms': Arthritis, arthralgia in multiple joints
- 'renal_involvement': Proteinuria, hematuria, or explicit mention of lupus nephritis Class II, III
, IV, or V
- 'serositis': Pleuritis, pericarditis
- 'dmards': Hydroxychloroquine, methotrexate, mycophenolate mofetil, azathioprine, belimumab, or
cyclophosphamide
- 'steroids': Corticosteroids (e.g., prednisone)

Provide a JSON object with '1' for present, '0' for absent, and '0.5' for suspected/unclear:

OUTPUT:
{{
  "{phenotype}": "1 or 0",
  "ANA": "1 or 0",
  "specific_antibodies": "1 or 0",
  "complement_levels": "1 or 0",
  "skin_manifestations": "1 or 0",
  "joint_symptoms": "1 or 0",
  "renal_involvement": "1 or 0",
  "serositis": "1 or 0",
  "dmards": "1 or 0",
  "steroids": "1 or 0"
}}
'''

PROMPT_ETP_METASTATIC_CANCER = '''
Analyze the provided discharge summary segment to determine the presence of {phenotype} and
associated details.
Use your clinical expertise to assess signs, symptoms, treatments, diagnostic criteria, and disease
 progression indicative of {phenotype}
in the context of solid tumor and not hematologic cancer.

Discharge Summary or Segment to Analyze:
{discharge_summary}

Instructions:
Determine if any of the following elements related to {phenotype} are present in the segment:
- '{phenotype}': Explicit mention of solid metastatic cancer, stage IV cancer, advanced cancer, or
terminology indicating spread beyond the primary site (e.g., "metastasized", "disseminated disease
", "distant spread")
- 'primary_tumor_site': Identification of the primary solid tumor location or origin of cancer (e.g
., "primary breast cancer", "lung adenocarcinoma", "colorectal tumor")
- 'metastasis_sites': Documented locations of metastasis, such as bones, liver, lungs, brain, lymph
 nodes, or any mention of distant organ involvement (e.g., "liver metastases", "bone lesions", "
brain mets")
- 'biopsy_results': Histological confirmation from biopsy of metastatic sites, including pathology
reports indicating metastatic disease (e.g., "biopsy-proven metastasis")
- 'oncological_treatments': Use of treatments specific to metastatic cancer, such as systemic
chemotherapy, targeted therapy, immunotherapy, or palliative radiotherapy (e.g., "started on
```

```
pembrolizumab", "palliative radiation to bone metastases")

Conclude with a JSON object that states the status of each predefined element. Respond with '1' for
 present and '0' for absent.

OUTPUT:
{{
  "{phenotype}": "1 or 0",
  "primary_tumor_site": "1 or 0",
  "metastasis_sites": "1 or 0",
  "biopsy_results": "1 or 0",
  "oncological_treatments": "1 or 0",
}}
'''

PROMPT_ETP_RHEUMATOID_ARTHRITIS = '''
Analyze the provided discharge summary segment to determine the presence of {phenotype} and
associated details.
Use your judgment to assess clinical signs, treatments, diagnostic criteria, and disease
progression indicative of {phenotype} seropositive or seronegative.

Discharge Summary or Segment to Analyze:
{discharge_summary}

Instructions:
Determine if any of the following elements related to {phenotype} are present in the segment:
- '{phenotype}': Explicit mention of rheumatoid arthritis, RA, or terminology indicating chronic
inflammatory joint disease (e.g., "rheumatoid arthritis", "RA", "inflammatory arthritis")
- 'joint_symptoms': Swelling, pain, stiffness in multiple joints, especially hands and feet
- 'morning_stiffness': Stiffness lasting >1 hour in the morning
- 'serological_markers': Rheumatoid factor (RF), anti-cyclic citrullinated peptide (anti-CCP)
antibodies
- 'imaging_findings': Joint erosions, narrowing of joint spaces on X-rays, MRI, or ultrasound
- 'cdmards': Use of conventional disease-modifying antirheumatic drugs (e.g., methotrexate,
sulfasalazine, leflunomide)
- 'biologics': Use of biologic agents (e.g., TNF inhibitors, rituximab, abatacept, tocilizumab, JAK
 inhibitors)
- 'steroids': Use of corticosteroids for RA management
- 'extra-articular_manifestations': Rheumatoid nodules, lung involvement, vasculitis

Conclude with a JSON object that states the status of each predefined element. Respond with '1' for
 present and '0' for absent.

OUTPUT:
{{
  "{phenotype}": "1 or 0",
  "joint_symptoms": "1 or 0",
  "morning_stiffness": "1 or 0",
  "serological_markers": "1 or 0",
  "imaging_findings": "1 or 0",
  "cdmards": "1 or 0",
  "biologics": "1 or 0",
  "steroids": "1 or 0",
  "extra-articular_manifestations": "1 or 0"
}}
'''

PROMPT_ETP_OBESITY = '''
```

```
Analyze the provided discharge summary segment to determine the presence of {phenotype} and
associated details.
Use your judgment to assess clinical signs, risk factors, treatments, and complications indicative
of {phenotype}.

Discharge Summary or Segment to Analyze:
{discharge_summary}

Instructions:
Determine if any of the following elements related to {phenotype} are present in the segment:
- '{phenotype}': Explicit mention of obesity or terminology indicating excessive body weight (e.g.,
 "morbid obesity", "severe obesity")
- 'BMI': Documented Body Mass Index (BMI) categorizing the patient as obese (BMI 30 or higher), if
not explicitly mentioned, use clinical judgment based on weight and height.
- 'waist_circumference': Measurement of waist circumference indicating abdominal obesity (greater
than 40 inches in men and 35 inches in women)
- 'comorbid_conditions': Presence of obesity-related comorbidities such as type 2 diabetes,
hypertension, sleep apnea, or dyslipidemia.
- 'bariatric_surgery': History of or planned bariatric surgery
- 'medication_for_obesity': Use of medications specifically aimed at weight loss (e.g., orlistat,
liraglutide, ozempic)

Conclude with a JSON object that states the status of each predefined element. Respond with '1' for
 present and '0' for absent.

OUTPUT:
{{
  "{phenotype}": "1 or 0",
  "BMI": "1 or 0",
  "waist_circumference": "1 or 0",
  "comorbid_conditions": "1 or 0",
  "bariatric_surgery": "1 or 0",
  "medication_for_obesity": "1 or 0"
}}
'''

PROMPT_ETP_ALCOHOL_ABUSE = '''
Analyze the provided discharge summary segment to determine the presence of alcohol abuse and
associated details.
Use your judgment to assess signs of substance abuse, treatments, and health consequences
indicative of alcohol abuse.

Discharge Summary or Segment to Analyze:
{discharge_summary}

Instructions:
Determine if any of the following elements related to {phenotype} are present in the segment:
- '{phenotype}': Explicit mention of alcohol abuse, alcohol use disorder, or alcoholism
- 'consumption_patterns': Heavy or frequent alcohol use, binge drinking
- 'withdrawal_symptoms': Signs of alcohol withdrawal such as tremors, agitation, or seizures.
- 'liver_abnormalities': abnormal liver enzymes, signs of alcoholic liver disease (labs or imaging)
- 'comorbid_conditions': Alcohol-related pancreatitis, gastritis, alcoholic liver disease
- 'treatment_for_addiction': Involvement in alcohol addiction treatment programs or counseling.
- 'medications': Use of disulfiram, naltrexone, acamprosate

Conclude with a JSON object that states the status of each predefined element. Respond with '1' for
 present and '0' for absent.

OUTPUT:
```

```
{{
  "{phenotype}": "1 or 0",
  "consumption_patterns": "1 or 0",
  "withdrawal_symptoms": "1 or 0",
  "liver_abnormalities": "1 or 0",
  "comorbid_conditions": "1 or 0",
  "treatment_for_addiction": "1 or 0",
  "medications": "1 or 0"
}}
'''

PROMPT_ETP_VENOUS_THROMBOEMBOLISM_PAST = '''
Analyze the provided discharge summary to determine any past medical history of venous
thromboembolism prior to the current admission.
Use your judgment to assess historical documentation of prior VTE episodes.

Discharge Summary to Analyze:
{discharge_summary}

Instructions:
Identify these historical VTE-related elements from before the current admission:
- '{phenotype}': Explicit mention of any VTE (DVT, PE, Splanchnic Venous Thrombosis or Cerebral
Venous Sinus Thrombosis)
- 'dvt_past': Prior deep vein thrombosis
- 'pe_past': Prior pulmonary embolism
- 'svt_past': Prior splanchnic venous thrombosis (portal, mesenteric, splenic, hepatic veins)
- 'cvst_past': Prior cerebral venous sinus thrombosis
- 'recurrent': Documentation of recurrent or multiple past episodes
- 'anticoagulation': Previous or active anticoagulation treatment for past history of VTE (heparin,
 low molecular weight heparin, warfarin, DOACs (e.g., apixaban, rivaroxaban)
- 'ivc_filter': History of IVC filter placement
- 'thrombophilia': Known thrombophilia or hypercoagulable condition

Conclude with a JSON object that states the status of each predefined element. Respond with '1' for
 present and '0' for absent.

OUTPUT:
{{
  "{phenotype}": "1 or 0",
  "dvt_past": "1 or 0",
  "pe_past": "1 or 0",
  "svt_past": "1 or 0",
  "cvst_past": "1 or 0",
  "recurrent": "1 or 0",
  "anticoagulation": "1 or 0",
  "ivc_filter": "1 or 0",
  "thrombophilia": "1 or 0"
}}
'''

PROMPT_ETP_VENOUS_THROMBOEMBOLISM_COMPLICATION = '''
Analyze the provided discharge summary to determine if venous thromboembolism occurred as a
complication during the current admission.
Use your judgment to assess new onset VTE during this hospitalization.

Discharge Summary to Analyze:
{discharge_summary}

Instructions:
```

```
Identify these VTE-related elements that developed during the current admission:
- '{phenotype}': New diagnosis of any VTE during admission (DVT, PE, Splanchnic Venous Thrombosis,
 orCerebral Venous Sinus Thrombosis)
- 'dvt_current': New DVT
- 'pe_current': New PE
- 'svt_current': New splanchnic venous thrombosis (portal, mesenteric, splenic, hepatic veins)
- 'cvst_current': New cerebral venous sinus thrombosis
- 'vte_symptoms': Clinical presentation (e.g., leg swelling, chest pain, abdominal pain, headache)
- 'diagnostic_tests': Positive imaging (ultrasound, CT, MRI/MRV, V/Q scan) or elevated D-dimer
- 'anticoagulation': Anticoagulation initiated for new VTE during current admission (heparin, low
molecular weight heparin, warfarin, DOACs (e.g., apixaban, rivaroxaban)
- 'interventions': Specific interventions for VTE (thrombolysis, thrombectomy, IVC filter)
- 'nosocomial': Clear evidence VTE developed >48 hours after admission

Conclude with a JSON object that states the status of each predefined element. Respond with '1' for
 present and '0' for absent.

OUTPUT:
{{
  "{phenotype}": "1 or 0",
  "dvt_current": "1 or 0",
  "pe_current": "1 or 0",
  "svt_current": "1 or 0",
  "cvst_current": "1 or 0",
  "vte_symptoms": "1 or 0",
  "diagnostic_tests": "1 or 0",
  "anticoagulation": "1 or 0",
  "interventions": "1 or 0",
  "nosocomial": "1 or 0",
}}
'''
```
